# Baseline Prognosis and Treatment Benefit From First-Line Immunotherapy Strategies in Advanced Solid Tumors: A Multicohort Target Trial Emulation

**DOI:** 10.64898/2026.11.20.26368610

**Authors:** Xavier Orcutt, Vivek Nimgaonkar, Kan Chen, Irbaz B. Riaz, Aaron B. Cohen, Ronac Mamtani, Manuel Gomes, Qi Long, Ravi B. Parikh

## Abstract

Background

In randomized trials, first-line immune checkpoint inhibitor (ICI)-based therapy has shown excess early mortality versus chemotherapy- or targeted therapy-containing regimens. ICI monotherapy is nonetheless often chosen for frail patients because it is perceived as less toxic, although these patients may be most vulnerable to this early mortality. Whether treatment benefit varies with baseline prognosis is unknown, as poor-prognosis patients are underrepresented in trials.

**Methods:** Using the US Flatiron Health electronic health record-derived database, we emulated target trials among 12,656 patients initiating first-line therapy during 2011–2023 across five comparisons: pembrolizumab plus chemotherapy versus pembrolizumab in head and neck squamous cell carcinoma (HNSCC; PD-L1 CPS ≥1 or unknown) and non–small cell lung cancer (NSCLC; PD-L1 TPS ≥50%); carboplatin plus gemcitabine versus pembrolizumab in urothelial carcinoma; pembrolizumab versus chemotherapy in dMMR/MSI-H colorectal cancer; and ipilimumab plus nivolumab versus BRAF/MEK inhibition in BRAF-mutant melanoma. Baseline prognosis was estimated with machine learning models predicting 6-month survival. Overlap-weighted pseudo-observation regression evaluated whether differences in 2-year restricted mean survival time (RMST) varied with predicted survival.

**Results:** In HNSCC, adding chemotherapy to pembrolizumab did not clearly improve RMST overall (11.2 days; 95% CI, -21.4 to 44.5), but benefit was 96.7 days greater at the 10th versus 90th percentile of predicted survival (95% CI, 33.7 to 159.7; interaction *P=*.003). Similar gradients favored carboplatin plus gemcitabine over pembrolizumab in urothelial carcinoma (10th–90th percentile difference, 51.4 days; 95% CI, 4.8 to 98.1; *P*=.03) and, less certainly, adding chemotherapy to pembrolizumab in NSCLC (35.3 days; 95% CI, -3.2 to 73.8; *P*=.07). Higher-risk patients in these cohorts were more often trial-ineligible. ICI-based therapy improved overall RMST in colorectal cancer (difference, 79.7 days; 95% CI, 49.5 to 107.0) and melanoma (68.3 days; 95% CI, 33.9 to 101.9), without clear variation by predicted survival (interaction *P*=.32 and *P*=.91).

**Conclusions:** Among patients with poorer prognosis, chemotherapy-containing regimens were associated with greater benefit than ICI monotherapy in HNSCC and urothelial carcinoma, and possibly NSCLC. ICI-based therapy appeared beneficial across prognosis in dMMR/MSI-H colorectal cancer and BRAF-mutant melanoma. Baseline prognosis may complement tumor biomarkers when selecting first-line immunotherapy strategies.

## Introduction

Immune checkpoint inhibitors (ICIs) have transformed the treatment of advanced solid tumors, enabling a subset of patients to achieve long-term survival rarely observed with chemotherapy [1]. Across multiple cancer types, randomized controlled trials (RCTs) have established ICIs as a standard component of first-line therapy, often in combination with chemotherapy [2–5]. While ICIs can produce durable, long-term benefit in a subset of patients, many patients derive limited or no benefit [6]. As ICIs have become embedded in routine practice, a central question is which patients benefit most relative to available alternatives.

Current approaches to selecting ICI therapy rely primarily on biomarkers of immune responsiveness, such as PD-L1 expression, tumor mutational burden, and mismatch repair status [7]. These markers, however, capture only one dimension of treatment selection. In many landmark RCTs, including those enriched for favorable biomarker profiles, ICI monotherapy is associated with higher mortality during the first few months of treatment compared with chemotherapy or targeted therapy containing regimens, followed by later divergence in favor of ICI [8–11].

Patients with lower baseline predicted survival may be vulnerable during this early period, when differences in the timing or probability of disease control could outweigh the potential for more durable ICI benefit. Yet the implications of baseline predicted survival for the choice between ICI and alternative therapies remain poorly characterized. We hypothesized that patients with lower baseline predicted survival may derive greater benefit from therapies providing more reliable early disease control, such as chemotherapy or targeted therapy, relative to ICI monotherapy.

RCTs are often limited in evaluating treatment-effect heterogeneity across baseline prognosis because patients with poor prognosis may be underrepresented and subgroup analyses are typically underpowered to detect effect modification [12,13]. Large, longitudinal oncology electronic health record (EHR) databases offer opportunities to evaluate this heterogeneity in routine practice [14]. Target trial emulation provides a framework for designing comparative effectiveness analyses using these observational data [15]. At the same time, predictive approaches to treatment-effect heterogeneity have shifted the focus from one-variable-at-a-time subgroup analyses toward evaluating treatment benefit according to a patient’s multivariable baseline risk [16]. Machine learning (ML) methods can estimate this risk from routinely collected EHR data [17,18].

We used target trial emulation to evaluate five first-line comparisons of ICI alone versus regimens including chemotherapy or targeted therapy across advanced solid tumors. We assessed whether treatment benefit varied with predicted survival to inform treatment selection, using ML prognostic models to estimate survival at treatment initiation.

## Methods

### Data Source

This study was a multi-cohort observational analysis using the EHR-derived, deidentified Flatiron Health Research Database. First-line treatment-initiation windows ranged from 2011 to 2023. The database contains longitudinal clinical data from approximately 220 U.S. cancer clinics, predominantly community oncology practices. It includes structured data (e.g., laboratory values, ICD codes) and unstructured data curated via technology-enabled abstraction of clinical notes and pathology reports by trained abstractors. Mortality was ascertained using a composite endpoint integrating multiple data sources that has demonstrated high concordance with the National Death Index [19].

### Study Cohorts

We identified first-line treatment comparisons involving ICI-based therapy and chemotherapy, targeted therapy, or combination regimens across advanced solid tumors. Comparisons were selected based on their relevance to the study question, available follow-up, and a pragmatic minimum of 300 patients per treatment arm to limit the inclusion of comparisons with sparse treatment groups. Five comparisons met these criteria:

1. Recurrent or metastatic head and neck squamous cell carcinoma (HNSCC) with PD-L1 combined positive score (CPS) ≥1 or unknown: pembrolizumab plus platinum-based chemotherapy versus pembrolizumab monotherapy.
2. Advanced non–small cell lung cancer (NSCLC) with PD-L1 tumor proportion score (TPS) ≥50%: pembrolizumab plus platinum-based chemotherapy versus pembrolizumab monotherapy.
3. Advanced urothelial carcinoma: carboplatin plus gemcitabine versus pembrolizumab.
4. Metastatic mismatch repair–deficient or microsatellite instability–high (dMMR/MSI-H) colorectal cancer: pembrolizumab versus fluoropyrimidine-based combination chemotherapy, with or without biologic therapy.
5. Advanced BRAF-mutant melanoma: ipilimumab plus nivolumab versus BRAF/MEK inhibitor combination therapy.

Each cohort was designed using target trial emulation principles and reported in accordance with the TARGET guideline [20] (Table S1); target trial specifications for each cohort are provided in Tables S2–S7. Eligible patients were new initiators of one of the treatment strategies of interest, with time zero defined as initiation of first-line systemic therapy for advanced or metastatic disease. Patients were analyzed according to their initial treatment strategy regardless of subsequent treatment changes, consistent with an intention-to-treat analogue. Treatment strategies and biomarker eligibility criteria were informed by analogous RCTs, including KEYNOTE-048 (HNSCC) [3], PERSEE (NSCLC) [21], KEYNOTE-361 (urothelial carcinoma) [22], KEYNOTE-177 (colorectal cancer) [8], and DREAMseq (melanoma) [10]. Laboratory and performance-status eligibility criteria were broader than those of the corresponding trials to capture the real-world spectrum of baseline predicted survival. Candidate treatment comparisons and their inclusion status are summarized in Table S8, cohort-specific treatment-initiation windows and data-lock dates in Table S9, and cohort flow diagrams in Figure S1.

### Estimation of Baseline Predicted Survival

For each cancer type, we developed a gradient-boosted survival model to predict 6-month survival from initiation of first-line therapy. The 6-month horizon was selected to characterize short-term prognosis when early differences between ICI and chemotherapy or targeted therapy are commonly observed [9,10,22]. Model-development populations included all patients with the corresponding advanced cancer initiating first-line systemic therapy, regardless of regimen or study-cohort eligibility.

Models used routinely collected demographic, tumor, vital-sign, laboratory, diagnostic-code, and medication features. Treatment assignment was excluded from the feature set. The resulting score represents an “observed-treatment” prognosis, reflecting survival under the treatment patterns in the model-development population rather than under a fixed reference treatment. Clinical features were measured at or before treatment initiation. For tumor biomarkers, results reported within 30 days after treatment initiation were permitted to account for delays between specimen collection and result availability. The influence of this window was examined in a sensitivity analysis. Insurance type, socioeconomic status, and treatment year were excluded from prognostic models but retained for confounding adjustment.

Each patient’s prediction was generated through cross-validation using a model not trained on that patient and calibrated using isotonic regression. Discrimination was assessed using the 6-month time-dependent area under the receiver operating characteristic curve (AUC) in held-out test data. Model-development populations, feature construction, missing-data handling, tuning, calibration, and performance are detailed in the Supplementary Methods (Table S10).

### Outcome and Estimand

The primary outcome was overall survival from first-line treatment initiation. The primary estimand was the between-group difference in 2-year restricted mean survival time (RMST). RMST measures expected survival accumulated through a specified horizon without requiring the proportional-hazards assumption, which may be inappropriate for ICI comparisons [23].

### Continuous Treatment-Effect Heterogeneity

To evaluate whether treatment benefit varied with baseline predicted survival, jackknife pseudo-observations were calculated for 2-year RMST [24]. These pseudo-observations were analyzed using an overlap-weighted least-squares regression model containing treatment assignment, calibrated predicted 6-month survival probability, and a treatment-by-predicted-survival interaction. The fitted model was used to estimate a continuous treatment-effect function representing the expected difference in 2-year RMST between treatment strategies at a calibrated 6-month survival probability (*r*).

Evidence that treatment effects varied with baseline predicted survival was assessed using the treatment-by-predicted-survival interaction coefficient, with statistical significance evaluated using a two-sided Wald test with robust standard errors. To provide clinical context for the magnitude of treatment effects across the risk distribution, we prespecified a 30-day difference in RMST as a pragmatic benefit threshold.

### Risk-Stratified Outcome Estimation

For comparisons in which the treatment-effect function reached the prespecified 30-day RMST benefit threshold, the corresponding predicted 6-month survival probability (*r*^∗^) defined higher risk (*r* < *r*^∗^) and lower-risk (*r* ≥ *r*^∗^) strata. These strata summarize the continuous treatment-effect relationship rather than establish a clinical decision boundary. Within each stratum, stabilized inverse probability weights (IPTW) were re-estimated, and RMST differences were calculated from weighted Kaplan–Meier curves. Confidence intervals were obtained using 1,000 bootstrap resamples.

### Confounding Adjustment

Propensity scores were estimated separately within each study cohort using logistic regression. Models included demographic, clinical, cancer-specific, laboratory, and healthcare-access variables measured at or before treatment initiation (Table S11). Overlap weighting was used for the primary treatment-effect heterogeneity analysis. This approach emphasizes patients in the region of clinical equipoise who could reasonably have received either treatment and reduces reliance on extrapolation in regions of limited propensity-score overlap [25]. IPTW was used to estimate marginal survival curves and RMST differences in the full cohort and within risk-defined strata. Covariate balance was evaluated using standardized mean differences, with an absolute standardized mean difference below 0.10 considered evidence of adequate balance [26]. Propensity-score specifications, weighting diagnostics, and baseline variable missingness are detailed in the Supplementary Methods and Supplementary Results.

### Robustness Analyses

We compared overall treatment effects with available RCT benchmarks using significance, confidence-interval, and standardized-difference agreement criteria adapted from RCT-DUPLICATE [27]. Because KEYNOTE-048 did not directly compare pembrolizumab plus chemotherapy with pembrolizumab monotherapy, the HNSCC cohort was compared descriptively with the corresponding KEYNOTE-048 arms using median overall survival. We also evaluated whether the framework reproduced treatment-effect heterogeneity observed in CheckMate 214 by comparing ipilimumab plus nivolumab with single-agent VEGF-targeted therapy in metastatic renal cell carcinoma (RCC), including clear-cell and not otherwise specified (NOS) histology to retain sufficient sample size, using 3-year RMST [4].

Sensitivity analyses limited biomarker results to those available by treatment initiation; used ridge-penalized Cox prognostic models; trained prognostic models only in chemotherapy-treated patients; modeled treatment-effect heterogeneity using natural cubic splines; evaluated 1-year RMST; applied alternative benefit thresholds of 14 and 60 days; and restricted the HNSCC analysis to documented PD-L1 CPS ≥1.

## Results

### Study Cohorts and Prognostic Model Performance

The five study cohorts included 12,656 patients with advanced or metastatic HNSCC, NSCLC, urothelial carcinoma, colorectal cancer, or melanoma initiating one of the first-line treatment strategies of interest (Table 1). Cohort sizes ranged from 1,043 patients with dMMR/MSI-H colorectal cancer to 5,605 patients with NSCLC. Median follow-up ranged from 24.9 to 66.5 months across treatment arms, exceeding the prespecified 2-year RMST horizon. Among cohorts with biomarker-based eligibility criteria, HNSCC was the only one that permitted unknown biomarker status at treatment initiation: 603 of 1,471 patients (41.0%) had documented PD-L1 CPS ≥1, while 868 (59.0%) had unknown PD-L1 status.

**Table 1.**
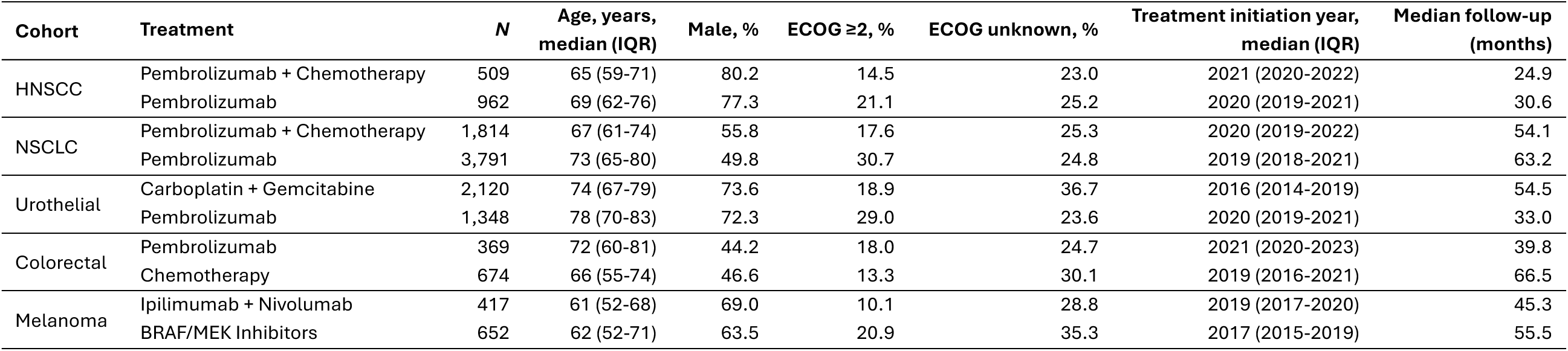
Patient characteristics and follow-up by cohort and treatment arm. This table summarizes unweighted patient characteristics and follow-up by treatment arm across five advanced solid tumor cohorts receiving first-line systemic therapy. Age and treatment-initiation year are reported as median (IQR). ECOG performance status was defined using the last recorded value at or before treatment initiation. ECOG ≥2 percentages exclude patients with unknown ECOG status. Median follow-up was estimated from treatment initiation using the reverse Kaplan–Meier method. ECOG = Eastern Cooperative Oncology Group; IQR = interquartile range.

| **Cohort** | **Treatment** | ***N*** | **Age, years, median (IQR)** | **Male, %** | **ECOG ≥2, %** | **ECOG unknown, %** | **Treatment initiation year, median (IQR)** | **Median follow-up (months)** |
|---|---|---|---|---|---|---|---|---|
| HNSCC | Pembrolizumab + Chemotherapy | 509 | 65 (59-71) | 80.2 | 14.5 | 23.0 | 2021 (2020-2022) | 24.9 |
|  | Pembrolizumab | 962 | 69 (62-76) | 77.3 | 21.1 | 25.2 | 2020 (2019-2021) | 30.6 |
| NSCLC | Pembrolizumab + Chemotherapy | 1,814 | 67 (61-74) | 55.8 | 17.6 | 25.3 | 2020 (2019-2022) | 54.1 |
|  | Pembrolizumab | 3,791 | 73 (65-80) | 49.8 | 30.7 | 24.8 | 2019 (2018-2021) | 63.2 |
| Urothelial | Carboplatin + Gemcitabine | 2,120 | 74 (67-79) | 73.6 | 18.9 | 36.7 | 2016 (2014-2019) | 54.5 |
|  | Pembrolizumab | 1,348 | 78 (70-83) | 72.3 | 29.0 | 23.6 | 2020 (2019-2021) | 33.0 |
| Colorectal | Pembrolizumab | 369 | 72 (60-81) | 44.2 | 18.0 | 24.7 | 2021 (2020-2023) | 39.8 |
|  | Chemotherapy | 674 | 66 (55-74) | 46.6 | 13.3 | 30.1 | 2019 (2016-2021) | 66.5 |
| Melanoma | Ipilimumab + Nivolumab | 417 | 61 (52-68) | 69.0 | 10.1 | 28.8 | 2019 (2017-2020) | 45.3 |
|  | BRAF/MEK Inhibitors | 652 | 62 (52-71) | 63.5 | 20.9 | 35.3 | 2017 (2015-2019) | 55.5 |

Prognostic model development populations are summarized in Table S12. Prognostic model discrimination was consistent across cancer types, with 6-month AUCs ranging from 0.751 in HNSCC to 0.798 in melanoma in held-out test data (Table S13). Predictions were well calibrated and captured substantial variation in baseline prognosis within each cohort (Figure S2). Propensity-score overlap and covariate balance were adequate after weighting (Figures S3 and S4). Arm-specific effective sample sizes after weighting and missingness of key baseline variables are reported in Tables S14 and S15.

### Treatment-Effect Heterogeneity Across Cohorts

Across the five study cohorts, two patterns emerged (Figure 1 and Table 2). HNSCC and urothelial carcinoma demonstrated a risk-dependent pattern in which the absolute RMST benefit of chemotherapy-containing strategies increased among patients with lower baseline predicted survival. NSCLC showed a directionally similar pattern, although evidence of effect modification was less certain. In contrast, in colorectal cancer and melanoma, point estimates favored ICI-based therapy across the baseline predicted survival distribution consistent with an apparent risk-invariant pattern.

**Figure 1.**
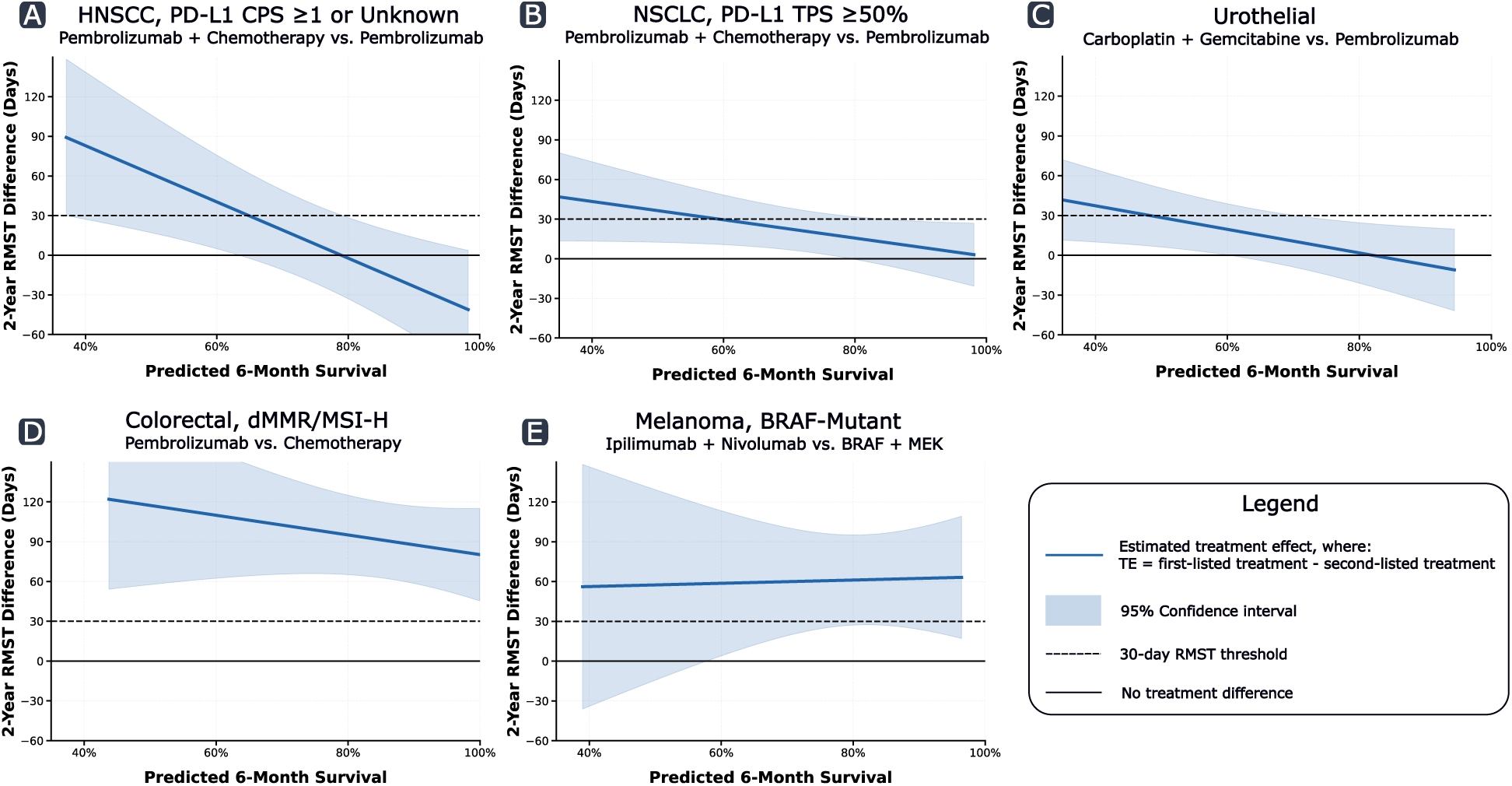
Heterogeneity of treatment effects according to baseline predicted survival across five advanced solid tumor cohorts. Estimated differences in 2-year restricted mean survival time (RMST) are shown as a function of calibrated predicted 6-month survival for (A) recurrent or metastatic head and neck squamous cell carcinoma (HNSCC) with PD-L1 combined positive score ≥1 or unknown, (B) advanced non-small cell lung cancer (NSCLC) with PD-L1 tumor proportion score ≥50%, (C) advanced urothelial carcinoma, (D) metastatic dMMR/MSI-H colorectal cancer, and (E) BRAF-mutant melanoma. Blue lines represent the estimated treatment-effect functions, and shaded areas represent 95% confidence intervals. Treatment effects were calculated as the first treatment listed in each panel minus the second. Thus, positive values favor the first treatment listed in each panel. Solid horizontal lines indicate no difference between treatments, and dashed horizontal lines indicate the prespecified 30-day RMST benefit threshold. Treatment-effect functions are displayed over the cohort-specific 5th to 95th percentiles of calibrated predicted 6-month survival.

**Table 2.** Overall treatment effects and treatment-effect heterogeneity across study cohorts. This table presents the overall inverse probability of treatment–weighted (IPTW) difference in 2-year restricted mean survival time (RMST) and estimates from the overlap-weighted continuous treatment-effect model. Positive RMST differences favor the first-listed treatment. Treatment effects (TEs) at the 10th and 90th percentiles represent the estimated differences in 2-year RMST evaluated at the overlap-weighted 10th and 90th percentiles of calibrated 6-month predicted survival, respectively. The 10th–90th difference is calculated as the TE at the 10th percentile minus the TE at the 90th percentile. A positive 10th–90th difference indicates greater absolute RMST benefit of that treatment among patients with worse baseline prognosis. Interaction *P* denotes the *P* value for the treatment-by-predicted-survival interaction. Values in parentheses are 95% confidence intervals. Pembro = Pembrolizumab; Chemo = Chemotherapy; Carbo/Gem = Carboplatin plus Gemcitabine; Ipi/Nivo = Ipilimumab plus Nivolumab.

| **Cohort** | **Comparison** | **Overall ΔRMST, days** | **TE at 10<sup>th</sup> percentile, days** | **TE at 90<sup>th</sup> percentile, days** | **10th–90th difference, days** | **Interaction *P*** |
|---|---|---|---|---|---|---|
| HNSCC | Pembro + Chemo vs. Pembro | 11.2 (-21.4 to 44.5) | 68.0 (20.3 to 115.8) | -28.6 (-67.7 to 10.4) | 96.7 (33.7 to 159.7) | .003 |
| NSCLC | Pembro + Chemo vs. Pembro | 24.4 (7.3 to 41.1) | 40.7 (13.1 to 68.3) | 5.4 (-16.6 to 27.4) | 35.3 (-3.2 to 73.8) | .07 |
| Urothelial | Carbo/Gem vs. Pembro | 23.2 (1.5 to 44.7) | 41.9 (11.5 to 72.3) | -9.6 (-39.3 to 20.2) | 51.4 (4.8 to 98.1) | .03 |
| Colorectal | Pembro vs. Chemo | 79.7 (49.5 to 107.0) | 108.4 (63.2 to 153.6) | 81.3 (47.5 to 115.0) | 27.2 (-25.9 to 80.2) | .32 |
| Melanoma | Ipi/Nivo vs. BRAF/MEK | 68.3 (33.9 to 101.9) | 56.8 (-25.1 to 138.7) | 63.0 (19.0 to 107.0) | -6.2 (-108.8 to 96.5) | .91 |

### Risk-Dependent Treatment Effects

In recurrent or metastatic HNSCC with PD-L1 CPS ≥1 or unknown, pembrolizumab plus chemotherapy was not associated with a statistically clear improvement in 2-year RMST compared with pembrolizumab monotherapy in the full cohort (difference, 11.2 days; 95% CI, −21.4 to 44.5), but the absolute RMST benefit increased as baseline predicted survival decreased (interaction *P* = .003; Figure 2). The estimated treatment effect differed by 96.7 days (95% CI, 33.7 to 159.7) between patients at the 10th and 90th percentiles of baseline predicted survival (Table 2). The treatment-effect function reached the prespecified 30-day RMST benefit threshold at a predicted 6-month survival probability of 0.65. Below this value, pembrolizumab plus chemotherapy was associated with a 94.3-day improvement in 2-year RMST (95% CI, 37.5 to 150.3) and higher 12-month overall survival (35.2% vs 20.2%), whereas no meaningful difference in 2-year RMST was observed above it (−12.7 days; 95% CI, −48.2 to 24.9; Table 3). In an analysis restricted to the 603 patients with documented PD-L1 CPS ≥1, the treatment-by-risk pattern was similar in direction and magnitude, although less precise (interaction *P* = .04; Table S30).

**Figure 2.**
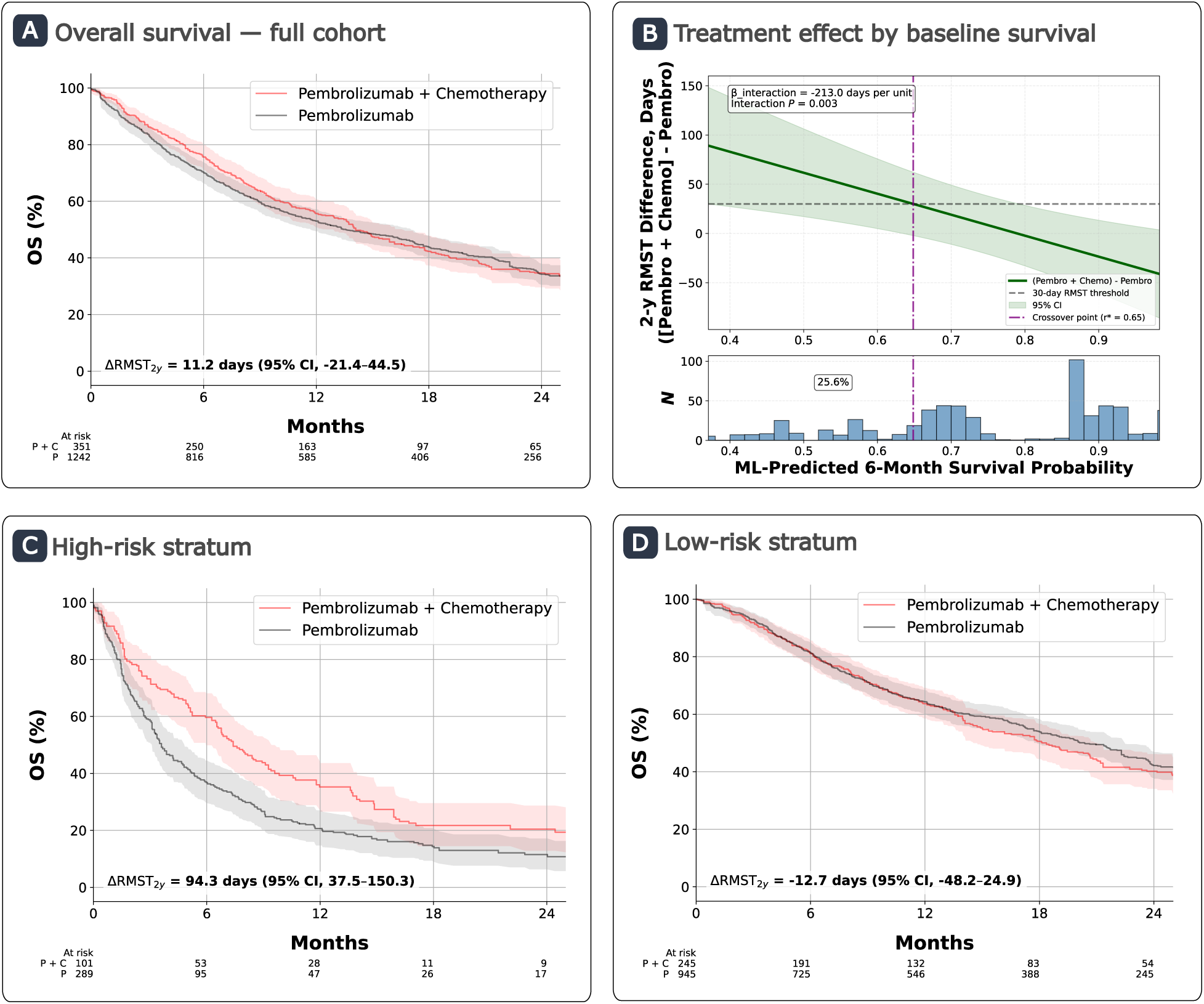
Heterogeneity of chemotherapy benefit by baseline predicted survival in R/M HNSCC with PD-L1 CPS ≥1 or unknown Panel. **A**: IPTW-adjusted overall survival in the full cohort, with no meaningful difference in 2-year RMST between pembrolizumab plus chemotherapy and pembrolizumab monotherapy (ΔRMST_2y =_ 11.2 days, 95% CI: -21.4 to 44.5). **Panel B**: Estimated treatment-effect function demonstrating increased benefit of combination therapy as baseline predicted survival worsened (β_interaction =_ −21.3 days per 0.1 increase in predicted 6-month survival, *P* = .003). The crossover point (r*), defined as the predicted survival probability at which combination therapy confers a ≥ 30-day improvement in 2-year RMST, occurred at r = 0.65. **Panels C and D**: IPTW-adjusted Kaplan–Meier curves stratified by r*. Among higher-risk patients (r < 0.65), combination therapy improved survival (ΔRMST_2y =_ 94.3 days, 95% CI: 37.5 to 150.3). Among lower-risk patients (r ≥ 0.65), no meaningful difference was observed (ΔRMST_2y =_ -12.7 days, 95% CI: -48.2 to 24.9). Numbers at risk represent IPTW-adjusted patient counts rather than raw patient counts.

**Table 3.**
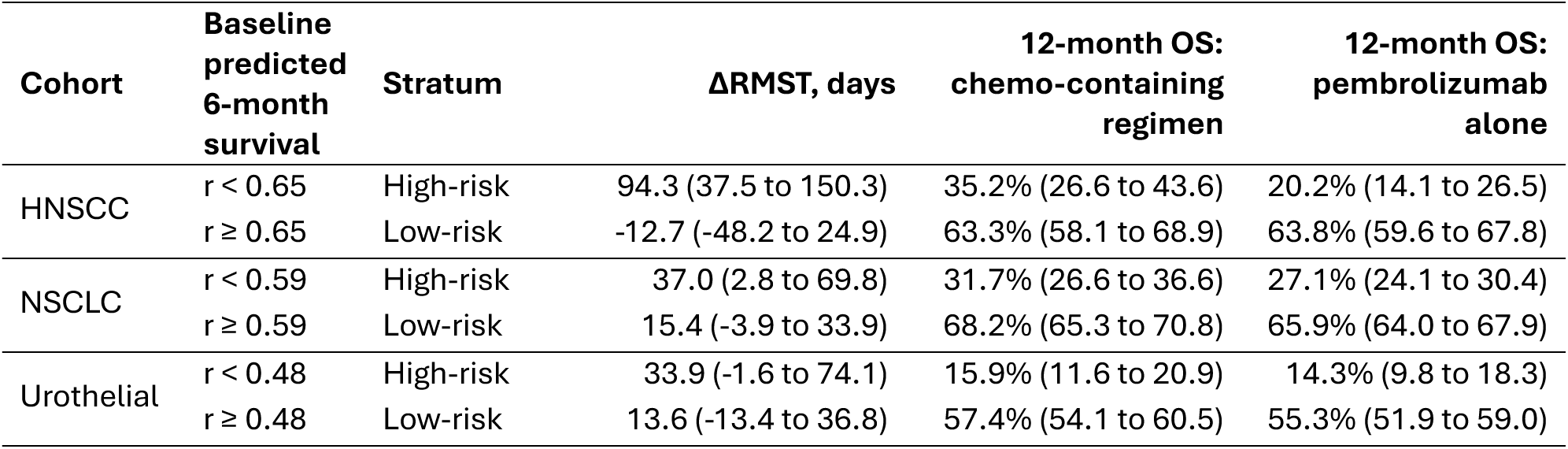
Risk-stratified treatment effects. This presents IPTW-adjusted RMST differences and 12-month overall survival by treatment within strata defined by calibrated baseline 6-month predicted survival (*r*). The cohort-specific cutoff is the value at which the estimated treatment-effect curve reaches the prespecified 30-day RMST benefit threshold; values below the cutoff define the higher-risk stratum. RMST differences use a 2-year horizon. The chemotherapy-containing regimen is pembrolizumab plus chemotherapy in HNSCC and NSCLC and carboplatin plus gemcitabine in urothelial carcinoma. Values in parentheses are 95% confidence intervals.

| Cohort | Baseline predicted 6-month survival | Stratum | ΔRMST, days | 12-month OS: chemo-containing regimen | 12-month OS: pembrolizumab alone |
|---|---|---|---|---|---|
| HNSCC | r < 0.65 | High-risk | 94.3 (37.5 to 150.3) | 35.2% (26.6 to 43.6) | 20.2% (14.1 to 26.5) |
|  | r ≥ 0.65 | Low-risk | -12.7 (-48.2 to 24.9) | 63.3% (58.1 to 68.9) | 63.8% (59.6 to 67.8) |
| NSCLC | r < 0.59 | High-risk | 37.0 (2.8 to 69.8) | 31.7% (26.6 to 36.6) | 27.1% (24.1 to 30.4) |
|  | r ≥ 0.59 | Low-risk | 15.4 (-3.9 to 33.9) | 68.2% (65.3 to 70.8) | 65.9% (64.0 to 67.9) |
| Urothelial | r < 0.48 | High-risk | 33.9 (-1.6 to 74.1) | 15.9% (11.6 to 20.9) | 14.3% (9.8 to 18.3) |
|  | r ≥ 0.48 | Low-risk | 13.6 (-13.4 to 36.8) | 57.4% (54.1 to 60.5) | 55.3% (51.9 to 59.0) |

Similar risk-dependent patterns were observed in the NSCLC and urothelial carcinoma study cohorts. In advanced NSCLC with PD-L1 TPS ≥50%, pembrolizumab plus chemotherapy was associated with a modest improvement in 2-year RMST compared with pembrolizumab monotherapy in the full cohort (difference, 24.4 days; 95% CI, 7.3 to 41.1), and the benefit increased as baseline predicted survival decreased, although the treatment-by-risk interaction did not reach statistical significance at the 2-year horizon (*P* = .07; Figure S5). The estimated treatment effect differed by 35.3 days (95% CI, -3.2 to 73.8) between patients at the 10th and 90th percentiles of baseline predicted survival (Table 2). Among patients with predicted 6-month survival below 0.59, pembrolizumab plus chemotherapy was associated with a 37.0-day improvement in 2-year RMST (95% CI, 2.8 to 69.8) and higher 12-month overall survival (31.7% vs 27.1%), whereas the estimated 2-year RMST benefit was smaller and not statistically significant among patients above this threshold (15.4 days; 95% CI, −3.9 to 33.9; Table 3).

In advanced urothelial carcinoma, carboplatin plus gemcitabine was associated with a modest improvement in 2-year RMST compared with pembrolizumab in the full cohort (difference, 23.2 days; 95% CI, 1.5 to 44.7), and its benefit increased as baseline predicted survival decreased (interaction *P* = .03; Figure S6). The estimated treatment effect differed by 51.4 days (95% CI, 4.8 to 98.1) between patients at the 10th and 90th percentiles of baseline predicted survival (Table 2), and the treatment-effect function reached the prespecified 30-day threshold at a predicted 6-month survival probability of 0.48. Below this value, the estimated 2-year RMST benefit of carboplatin plus gemcitabine was larger but imprecise (33.9 days; 95% CI, −1.6 to 74.1), and 12-month overall survival was similar between treatments (15.9% vs 14.3%). Above the threshold, the estimated benefit was smaller (13.6 days; 95% CI, −13.4 to 36.8; Table 3), and survival curves crossed after approximately 1 year, with a longer survival tail with pembrolizumab beyond the 2-year horizon (Figure S6D).

Higher-risk strata comprised 21.5% to 28.5% of patients in the HNSCC, NSCLC, and urothelial carcinoma cohorts. These patients had worse performance status, more frequent laboratory abnormalities, greater weight loss, and a higher burden of metastatic disease than lower-risk patients (Tables S16–S18).

### Apparent Risk-Invariant Treatment Effects

There was no clear evidence of risk-related treatment-effect modification in dMMR/MSI-H colorectal cancer or BRAF-mutant melanoma. Pembrolizumab improved 2-year RMST compared with chemotherapy in colorectal cancer (difference, 79.7 days; 95% CI, 49.5 to 107.0), while ipilimumab plus nivolumab improved 2-year RMST compared with BRAF/MEK inhibition in melanoma (difference, 68.3 days; 95% CI, 33.9 to 101.9). In both cohorts, point estimates were comparatively stable across baseline predicted survival, with no significant treatment-by-risk interaction or crossover of the prespecified 30-day RMST threshold (Figure 3 and Figure S7; Table 2). However, pointwise confidence intervals for the treatment-effect function were wide in melanoma, particularly at lower predicted survival probabilities. The estimated treatment-effect differences between the 10th and 90th percentiles were small in colorectal cancer (27.2 days; 95% CI, −25.9 to 80.2) and melanoma (−6.2 days; 95% CI, −108.8 to 96.5), although these estimates were imprecise (Table 2). In a flexible spline specification, the magnitude of pembrolizumab benefit in colorectal cancer varied across predicted survival (global *P* = .001), with the largest estimated benefit at intermediate predicted survival, but it remained above the 30-day threshold across the observed range and did not favor chemotherapy in any risk range (Table S27 and Figure S9).

**Figure 3.**
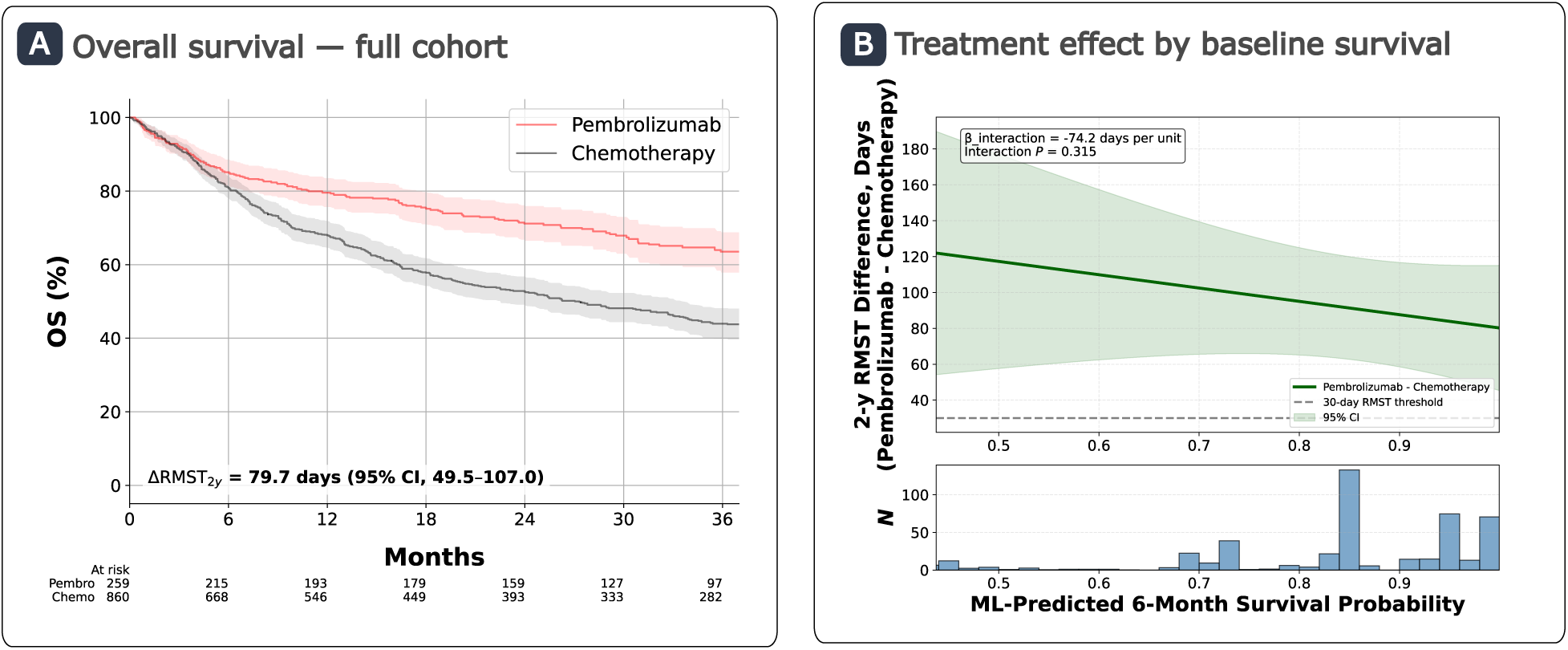
Heterogeneity of pembrolizumab benefit by baseline predicted survival in metastatic colorectal cancer with dMMR/MSI-H Panel. **A**: IPTW-adjusted overall survival in the full cohort demonstrating improved 2-year RMST with pembrolizumab compared with chemotherapy (ΔRMST_2y =_ 79.7 days, 95% CI: 49.5 to 107.0). Numbers at risk represent IPTW-adjusted patient counts rather than raw patient counts. **Panel B**: Estimated treatment-effect function demonstrated no clear evidence of a linear interaction across baseline predicted survival (β_interaction =_ -7.4 days per 0.1 increase in predicted 6-month survival; *P* = .32). No crossover point was identified.

### Robustness Analyses

Across four comparisons with suitable RCT benchmarks, all emulations met confidence-interval and standardized-difference agreement criteria, and two met significance agreement (Table S19). We next compared the three risk-dependent cohorts with their corresponding trials, because these were the comparisons in which an overall treatment effect could obscure differences across baseline risk. Overall treatment effects aligned with PERSEE in NSCLC and with KEYNOTE-361 in urothelial carcinoma, and median overall survival in HNSCC was similar to that in the corresponding KEYNOTE-048 arms, which were not directly compared in that trial (Table S20). In all three cohorts, higher-risk patients were more frequently classified as trial-ineligible based on performance-status or laboratory criteria than lower-risk patients (HNSCC, 22.4% vs 4.9%; NSCLC, 26.2% vs 5.1%; urothelial carcinoma, 56.0% vs 21.1%; Table S21).

In a secondary concordance analysis of metastatic RCC, ipilimumab plus nivolumab demonstrated greater benefit relative to single-agent VEGF-targeted therapy among patients with poorer baseline predicted survival, consistent with the treatment-effect heterogeneity observed in CheckMate 214 (Table S22 and Figure S8). The model-defined higher-risk group was strongly enriched for IMDC intermediate- or poor-risk disease (Table S23).

Treatment-effect patterns were generally preserved across sensitivity analyses. Detailed sensitivity and exploratory results are provided in the Supplementary Results (Tables S24–S32; Figure S9).

## Conclusions

Across five comparisons of first-line ICI-based treatment strategies, we identified two patterns of treatment benefit according to baseline prognosis: risk-dependent and apparent risk-invariant. In the HNSCC, NSCLC, and urothelial carcinoma study cohorts, treatment effects varied with baseline prognosis, with chemotherapy-containing strategies becoming more favorable among patients with poorer predicted survival. The treatment- by-risk interaction in NSCLC did not reach statistical significance at the primary 2-year horizon, but its direction was consistent across sensitivity analyses, and it was statistically significant at the secondary 1-year horizon and when biomarker eligibility was restricted to results available at treatment initiation (Tables S24 and S28). The stronger signal at 1 year is consistent with a benefit of adding chemotherapy that is concentrated in the early treatment period. Its direction also aligned with prior real-world evidence suggesting greater benefit from adding chemotherapy to pembrolizumab among patients at high risk of early progression [28].

In contrast, the benefit of pembrolizumab over chemotherapy in dMMR/MSI-H colorectal cancer and of ipilimumab plus nivolumab over BRAF/MEK inhibition in BRAF-mutant melanoma appeared broadly consistent across the baseline-risk distribution. In both comparisons, the ICI-based strategy was associated with a significant overall survival benefit in the full study population, whereas the overall differences between strategies were smaller in the risk-dependent comparisons. These archetypes should be interpreted as properties of the specific treatment comparisons evaluated rather than intrinsic features of the cancer types or biomarker-defined populations.

These findings may have practical implications for treatment selection. Biomarkers such as PD-L1 expression and mismatch-repair status characterize tumor biology and the likelihood of benefit from ICI. Baseline predicted survival, integrating disease burden, physiologic reserve, laboratory abnormalities, and comorbidity, may provide complementary information. This may be particularly relevant when ICI monotherapy is considered for a clinically frail patient because it is perceived as less toxic than combination therapy [29]. In the risk-dependent cohorts studied here, the estimated benefit of chemotherapy-containing treatment was concentrated among patients with lower predicted survival. Although toxicity and treatment feasibility remain essential considerations, these findings suggest that selecting a less intensive regimen based primarily on perceived tolerability may not always be associated with better survival outcomes. Distinguishing whether poor prognosis is driven primarily by disease burden, which chemotherapy may address, or by comorbidity and limited physiologic reserve may further refine treatment selection.

In the risk-dependent cohorts, overall treatment effects were consistent with those of the corresponding trials, yet this agreement masked heterogeneity across baseline risk. Higher-risk patients, in whom chemotherapy-containing strategies were associated with the greatest benefit, were also those most often ineligible for these trials by performance-status or laboratory criteria. Trial results may therefore generalize well to the average real-world patient while providing limited guidance for higher-risk patients, in whom the choice between strategies was associated with the largest differences in survival. In this setting, risk-stratified analyses of real-world data may complement trial evidence by characterizing treatment effects in populations that trials underrepresent.

Several limitations should be considered. First, residual confounding remains possible despite propensity score adjustment [30]. Second, risk-based approaches to treatment-effect heterogeneity require caution when average treatment effects are small or null [31], although the risk-dependent findings persisted across alternative prognostic models and functional-form specifications. Third, predicted survival estimates represent an “observed-treatment” prognosis and may therefore reflect associations between baseline characteristics and treatment selection, rather than treatment-independent baseline risk. However, chemotherapy-only model development preserved the patterns in HNSCC and colorectal cancer cohorts. Fourth, PD-L1 status was unknown at treatment initiation for 59.0% of patients in the HNSCC cohort, although findings were similar among patients with documented CPS ≥1. Fifth, predicted 6-month survival probabilities in the colorectal cancer and melanoma cohorts were concentrated toward higher values, limiting information about treatment-effect heterogeneity among patients with poor predicted survival. Apparent risk invariance therefore requires cautious interpretation. Finally, crossover thresholds should be interpreted as descriptive summaries of continuous treatment-effect relationships rather than precise clinical decision boundaries.

In conclusion, the relationship between baseline prognosis and benefit from first-line immunotherapy strategies differed by treatment comparison. In HNSCC and urothelial carcinoma, and possibly NSCLC, chemotherapy-containing regimens were associated with greater survival benefit than pembrolizumab monotherapy among patients with poorer prognosis, a population underrepresented in the trials that established these regimens. In contrast, the benefit of ICI-based therapy in dMMR/MSI-H colorectal cancer and BRAF-mutant melanoma appeared consistent across prognosis. These findings suggest that baseline prognosis may complement tumor biomarkers when selecting first-line immunotherapy strategies, and they warrant evaluation with independently validated prognostic models.

## Declarations

### Ethics approval and consent to participate

This study was approved by the Emory University Institutional Review Board, which granted a waiver of informed consent because the data were deidentified and retrospective.

### Consent for publication

Not applicable.

### Availability of data and material

The Flatiron data used in this study were made available as part of an academic collaboration with Emory University. The deidentified data are subject to licensing restrictions and may be made available to qualified researchers through a proposal-based process. Requests should be directed to.

Code for data processing, model development, and statistical analyses is available at https://github.com/xavier-orcutt/io-heterogeneity.

### Competing interests

R.B.P. reports grants from the National Institutes of Health, Department of Defense, Prostate Cancer Foundation, NCCN Foundation, Humana, Emerson Collective, Schmidt Futures, Arnold Ventures, Mendel.ai, and Veterans Health Administration; personal fees and equity from GNS Healthcare, Thyme Care, Main Street Health, and Onc.AI; personal fees from ConcertAI, Cancer Study Group, Mendel.ai, Optinosis, Biofourmis, Archetype Therapeutics, CreditSuisse, G1 Therapeutics, Humana, and Nanology; stock options in Counsel Health and Scaled Insights; honoraria from Flatiron and Medscape; has board membership (unpaid) at the Coalition to Transform Advanced Care and American Cancer Society; editor at the Journal of Clinical Oncology; and served on a leadership consortium (unpaid) at the National Quality Forum, all outside the submitted work. V. N. reports personal fees and equity from Valar Labs, outside the submitted work. A.B.C. is an employee of Flatiron Health. All other authors declare no conflicts of interest.

### Funding

This work was supported by the following grants from the National Institutes of Health: K08CA263541 (R.B.P.), P30CA016520 (Q.L. and R.M.), U01CA274576 (Q.L.), and 5T32CA009071 (V.N.). The funders had no role in the design of the study; the collection, analysis, and interpretation of the data; the writing of the manuscript; or the decision to submit the manuscript for publication.

### Authors’ contributions

X.O. and V.N. contributed equally. Conceptualization: X.O., V.N., R.B.P. Methodology: X.O., V.N., K.C., Q.L., R.B.P. Formal analysis: X.O., V.N. Interpretation of data: all authors. Writing – original draft: X.O., V.N., R.B.P. Writing – review and editing: all authors. Supervision: Q.L., R.B.P. All authors approved the final manuscript. R.B.P. is the guarantor of the study.

## Acknowledgements

We thank Joshua Wallach of Emory University for their feedback on initial versions of the manuscript.

## Notes

### Clinical Protocols

https://doi.org/10.5281/zenodo.23113983

### Author Declarations

The Institutional Review Board of Emory University waived the requirement for informed consent because the study used deidentified, retrospective data.

### Summary of Updates

Version 3 updates PD-L1 ascertainment to use documented TPS and CPS values for patients with indeterminate abstracted biomarker status, who were previously classified as unknown, which increased the NSCLC cohort (n=5,605) and the proportion of HNSCC patients with documented CPS ≥1. All analyses were rerun. Main findings are unchanged; the NSCLC treatment-by-risk gradient was smaller with a similar P value (.07 vs .06).

