## supplement for "Baseline Prognosis and Treatment Benefit From First-Line Immunotherapy Strategies in Advanced Solid Tumors: A Multicohort Target Trial Emulation"

### Supplementary Appendix

#### Supplementary Methods

##### Reporting guideline and target trial specifications

This study was designed and reported in accordance with the Transparent Reporting of Observational Studies Emulating a Target Trial (TARGET) guideline [1]. The completed TARGET checklist is provided in Table S1, and target trial specifications for each study cohort are provided in Tables S2–S7.

**Table S1. TARGET Reporting Checklist**

| Item | Location in manuscript |
|---|---|
| **Abstract** | |
| 1a. Study identification and objectives | Title; Abstract: Background and Methods |
| 1b. Data sources | Abstract: Methods |
| 1c. Assumptions, methods, findings, conclusions | Abstract: Methods, Results, and Conclusions |
| **Introduction** | |
| 2. Background | Introduction: paragraphs 1-4 |
| 3. Causal question | Introduction: paragraphs 3 and 5; Tables S2-S7 |
| 4. Rationale | Introduction: paragraphs 1-4 |
| **Methods** | |
| 5. Data sources | Methods: Data Source |
| *6. Target trial specification* | |
| 6a. Eligibility criteria | Tables S2–S7 |
| 6b. Treatment strategies | Tables S2–S7 |
| 6c. Assignment procedures | Tables S2–S7 |
| 6d. Follow-up | Tables S2–S7 |
| 6e. Outcomes | Tables S2–S7 |
| 6f. Causal contrasts | Tables S2–S7 |
| 6g. Identifying assumptions | Tables S2–S7 |
| 6h. Data analysis plan | Tables S2–S7 |
| *7. Target trial emulation* | |
| 7a. Eligibility criteria | Methods: Study Cohorts; Tables S2–S7; Figure S1 |
| 7b. Treatment strategies | Methods: Study Cohorts; Tables S2–S7 |
| 7c. Assignment procedures | Methods: Confounding Adjustment; Tables S2–S7 and S11 |
| 7d. Follow-up | Methods: Study Cohorts; Outcome and Estimand; Tables S2–S7 and S9 |
| 7e. Outcomes | Methods: Data Source; Outcome and Estimand; Tables S2–S7 |
| 7f. Causal contrasts | Methods: Study Cohorts; Outcome and Estimand; Tables S2–S7 |
| 7g. Identifying assumptions | Methods: Confounding Adjustment; Tables S2–S7; Discussion: paragraph 5 |
| 7h. Data analysis plan | Methods: Estimation of Baseline Predicted Survival; Continuous Treatment-Effect Heterogeneity; Risk-Stratified Outcome Estimation; Confounding Adjustment; Robustness Analyses; Tables S2–S7 |
| **Results** | |
| 8. Participant selection | Results: Study Cohorts and Prognostic Model Performance; Table 1; Figure S1 |

| | |
|---|---|
| 9. Baseline data | Results: Study Cohorts and Prognostic Model Performance; Table 1 |
| 10. Follow-up | Results: Study Cohorts and Prognostic Model Performance; Tables 1 and S9 |
| 11. Missing data | Supplementary Methods: Feature construction and missing data handling; Table S15 |
| 12. Outcomes | Tables 2–3; Figures 1–3; Figures S5–S8 |
| 13. Effect estimates | Results: Risk-Dependent Treatment Effects; Apparent Risk-Invariant Treatment Effects; Tables 2–3; Figures 1–3 |
| 14. Additional analyses | Results: Robustness Analyses; Supplementary Results; Tables S19–S32 |
| **Discussion** | |
| 15. Interpretation | Conclusions: paragraphs 1-4 |
| 16. Limitations | Conclusions: paragraph 5 |
| **Other information** | |
| 17. Ethics | Declarations: Ethics approval and consent to participate |
| 18. Registration | Supplementary Methods: Trial protocol registration |
| 19. Sharing of study materials | Declarations: Availability of data and material; Supplementary Methods: Trial protocol registration |
| 20. Funding sources | Declarations: Funding |
| 21. Conflicts of interest | Declarations: Competing interests |

**Table S2. Target trial emulation specification for first-line pembrolizumab plus chemotherapy vs. pembrolizumab monotherapy in recurrent/metastatic head and neck squamous cell carcinoma with PD-L1 CPS ≥1 or unknown**

| TARGET component | Target trial specification | Emulation using observational data |
|---|---|---|
| Causal question | Primary causal question: among patients with recurrent/metastatic head and neck squamous cell carcinoma (R/M HNSCC) and positive or unknown PD-L1 initiating first-line therapy, how does the causal effect of pembrolizumab plus platinum-based chemotherapy versus pembrolizumab monotherapy on 2-year restricted mean survival time (RMST) vary with baseline predicted 6 month survival? | Same causal question addressed using the Flatiron Health Research Database. |
| Eligibility criteria | • R/M HNSCC<br>• Age ≥18<br>• PD-L1 combined positive score is ≥1 or unknown<br>• Initiating first-line systemic therapy for recurrent or metastatic disease<br>• Eligibility is pragmatic and does not include strict laboratory or comorbidity exclusions | Eligibility was assessed at time zero (first-line treatment initiation) using diagnosis, staging, treatment, and biomarker data recorded in the Flatiron Health Research Database.<br><br>PD-L1 tests reported up to 30 days after time zero were included, to allow for reporting delays. A test was considered positive if the abstracted status was positive or the documented CPS was ≥1, and negative if the status was negative or the CPS was <1. Tests with an indeterminate status but a documented CPS were classified by CPS. Patients were classified as positive if any test was positive, negative if all classifiable tests were negative, and unknown otherwise. Patients classified as negative were excluded.<br><br>Patients initiating treatment too close to the data cutoff to permit adequate follow-up for the prespecified RMST horizon were excluded. |
| Treatment strategies | **Strategy A:** Initiate pembrolizumab plus platinum-based chemotherapy (cisplatin- or carboplatin-based). | Treatment strategies were defined by the first-line systemic regimen recorded in the Flatiron Health Research Database. |

| | | |
|---|---|---|
| | **Strategy B:** Initiate pembrolizumab monotherapy. | **Strategy A:** Pembrolizumab plus platinum-based chemotherapy (cisplatin- or carboplatin-based).<br><br>**Strategy B:** Pembrolizumab monotherapy.<br><br>Regimens including targeted therapies, clinical study drugs, or immune checkpoint inhibitors other than pembrolizumab are excluded. |
| Assignment procedures | Eligible patients are randomly assigned 1:1 to Strategy A versus Strategy B at time zero. Treatment initiation occurs immediately after assignment. | Treatment assignment was defined by the first-line regimen initiated at time zero, as recorded in the Flatiron Health Research Database. In the absence of randomization, analyses estimated a per-assignment (intention-to-treat [ITT] analogue) effect of treatment initiation without censoring for subsequent treatment discontinuation or switching. Confounding was addressed using propensity score methods to balance baseline covariates, with overlap weighting used for treatment-effect heterogeneity analyses and stabilized inverse probability of treatment weighting (IPTW) used for Kaplan–Meier and RMST estimation in the full cohort and within risk-defined strata. |
| Follow-up | Start: Time of randomization and treatment initiation (time zero).<br><br>End: Death, loss to follow-up, or administrative censoring at the planned end of the study period. | Start: Time of first-line systemic therapy initiation, defined as the earliest point at which eligibility and treatment assignment are simultaneously established.<br><br>End: Death or censoring at last recorded activity in the EHR. |
| Outcomes | Primary outcome: Overall survival from time zero.<br>Primary summary measure: RMST through 2 years from time zero. | Primary outcome: Overall survival from time zero.<br><br>Primary summary measure: RMST through 2 years from time zero. |
| Causal contrasts | Primary contrast: Difference in 2-year RMST under assignment to Strategy A versus Strategy B. | Primary contrast: Marginal difference in 2-year RMST between treatment strategies in the full cohort, estimated |

| | | |
|---|---|---|
| | Heterogeneity analysis: Estimate the treatment-effect function TE(r) across baseline calibrated 6-month survival probability r using a treatment-by-risk interaction.<br><br>Risk stratification: Define a crossover point r* summarizing TE(r) at a clinically meaningful benefit threshold (30 days of 2-year RMST) and estimate treatment effects within r < r* and r≥ r* strata. | using stabilized IPTW.<br><br>Heterogeneity analysis: Treatment-effect heterogeneity was evaluated by modeling RMST as a function of baseline predicted 6-month survival probability, including a treatment-by-risk interaction term in an overlap-weighted regression model.<br><br>Risk stratification: A crossover point r* was defined as the value of baseline predicted survival at which the estimated treatment effect reached a prespecified threshold of 30 days. Treatment effects were then estimated within strata defined by r < r* and r ≥ r* using stabilized IPTW-adjusted Kaplan–Meier curves and RMST. |
| Identifying assumptions | Randomization at treatment initiation ensures exchangeability, consistency, and positivity by design. Censoring is assumed to be non-informative. | We assume conditional exchangeability of treatment initiation given measured baseline covariates, consistency of treatment strategies, positivity within the region of clinical equipoise emphasized by overlap weighting, and non-informative censoring conditional on baseline covariates. |
| Data analysis plan | (1) Measure baseline covariates at or before time zero.<br><br>(2) Develop and calibrate a prognostic model to predict 6-month survival from baseline covariates, excluding treatment assignment.<br><br>(3) Estimate the effect of assignment to Strategy A versus Strategy B on 2-year RMST using an ITT analysis.<br><br>(4) Evaluate absolute treatment-effect heterogeneity by modeling RMST as a function of baseline predicted 6-month survival probability, including a treatment-by-risk interaction. Estimate treatment | (1) Measure baseline covariates at or before time zero.<br><br>(2) Develop and calibrate a prognostic model to predict 6-month survival from baseline covariates, excluding treatment assignment.<br><br>(3) Estimate the marginal effect of Strategy A versus Strategy B on 2-year RMST using stabilized IPTW.<br><br>(4) Evaluate absolute treatment-effect heterogeneity by modeling RMST as a function of baseline predicted 6-month survival probability, including a treatment-by-risk interaction, using overlap-weighted regression. Estimate |

| | | |
|---|---|---|
| | effect function TE(r) and corresponding confidence intervals.<br><br>(5) Summarize TE(r) using a prespecified crossover point r*, defined as the value of baseline predicted survival at which the estimated treatment effect reaches 30 days of 2-year RMST. Report ITT effects within r < r* and r ≥ r* strata. | treatment effect function TE(r) and corresponding confidence intervals.<br><br>(5) Summarize TE(r) using a prespecified crossover point r*, defined as the value of baseline predicted survival at which the estimated treatment effect reaches 30 days of 2-year RMST. Estimate IPTW-adjusted survival and RMST within r < r* and r ≥ r* strata.<br><br>(6) Conduct sensitivity analyses to evaluate robustness to key analytic choices, including restriction to biomarker results available by treatment initiation, use of a ridge-penalized Cox prognostic model, prognostic model development restricted to chemotherapy-treated patients, a natural cubic spline treatment-effect specification, a 1-year RMST horizon, alternative clinically meaningful RMST benefit thresholds of 14 and 60 days, and cohort restriction to documented PD-L1 CPS ≥1. |

**Table S3. Target trial emulation specifications for first-line pembrolizumab plus chemotherapy vs. pembrolizumab monotherapy in PD-L1 TPS ≥50% advanced non-small cell lung cancer**

| TARGET component | Target trial specification | Emulation using observational data |
|---|---|---|
| Causal question | Primary causal question: among patients with advanced non-small cell lung cancer (NSCLC) and PD-L1 TPS ≥50% initiating first-line therapy, how does the causal effect of pembrolizumab plus platinum-based chemotherapy versus pembrolizumab monotherapy on 2-year restricted mean survival time (RMST) vary with baseline predicted 6-month survival probability? | Same causal question addressed using the Flatiron Health Research Database. |
| Eligibility criteria | • Advanced or metastatic NSCLC<br>• Age ≥18<br>• PD-L1 tumor proportion score ≥50%<br>• EGFR or ALK negative or unknown<br>• Initiating first-line systemic therapy for advanced disease<br>• Eligibility is pragmatic and does not include strict laboratory or comorbidity exclusions | Eligibility was assessed at time zero (first-line treatment initiation) using diagnosis, staging, treatment, and biomarker data recorded in the Flatiron Health Research Database.<br><br>PD-L1 tests reported up to 30 days after time zero were included, to allow for reporting delays. Eligibility was based on the highest documented TPS across tests, regardless of abstracted status. Patients with TPS ≥50% were eligible. Patients with TPS <50% or no documented TPS were excluded. Patients with any EGFR mutation or ALK rearrangement reported in the same window were excluded.<br><br>Patients initiating treatment too close to the data cutoff to permit adequate follow-up for the prespecified RMST horizon were excluded. |
| Treatment strategies | **Strategy A:** Initiate pembrolizumab plus platinum-based chemotherapy (cisplatin- or carboplatin-based).<br><br>**Strategy B:** Initiate pembrolizumab monotherapy. | Treatment strategies were defined by the first-line systemic regimen recorded in the Flatiron Health Research Database.<br><br>**Strategy A:** Pembrolizumab plus platinum-based chemotherapy (cisplatin- or carboplatin-based). |

| | | |
|---|---|---|
| | | **Strategy B:** Pembrolizumab monotherapy.<br><br>Regimens including targeted therapies, clinical study drugs, or immune checkpoint inhibitors other than pembrolizumab are excluded. |
| Assignment procedures | Eligible patients are randomly assigned 1:1 to Strategy A versus Strategy B at time zero. Treatment initiation occurs immediately after assignment. | Treatment assignment was defined by the first-line regimen initiated at time zero, as recorded in the Flatiron Health Research Database. In the absence of randomization, analyses estimated a per-assignment (intention-to-treat [ITT] analogue) effect of treatment initiation without censoring for subsequent treatment discontinuation or switching. Confounding was addressed using propensity score methods to balance baseline covariates, with overlap weighting used for treatment-effect heterogeneity analyses and stabilized inverse probability of treatment weighting (IPTW) used for Kaplan–Meier and RMST estimation in the full cohort and within risk-defined strata. |
| Follow-up | Start: Time of randomization and treatment initiation (time zero).<br><br>End: Death, loss to follow-up, or administrative censoring at the planned end of the study period. | Start: Time of first-line systemic therapy initiation, defined as the earliest point at which eligibility and treatment assignment are simultaneously established.<br><br>End: Death or censoring at last recorded activity in the EHR. |
| Outcomes | Primary outcome: Overall survival from time zero.<br><br>Primary summary measure: RMST through 2 years from time zero. | Primary outcome: Overall survival from time zero.<br><br>Primary summary measure: RMST through 2 years from time zero. |
| Causal contrasts | Primary contrast: Difference in 2-year RMST under assignment to Strategy A versus Strategy B.<br><br>Heterogeneity analysis: Estimate the treatment-effect function TE(r) across | Primary contrast: Marginal difference in 2-year RMST between treatment strategies in the full cohort, estimated using stabilized IPTW.<br><br>Heterogeneity analysis: Treatment- |

| | | |
|---|---|---|
| | baseline calibrated 6-month survival probability r using a treatment-by-risk interaction.<br><br>Risk stratification: Define a crossover point r* summarizing TE(r) at a clinically meaningful benefit threshold (30 days of 2-year RMST) and estimate treatment effects within r < r* and r≥ r* strata. | effect heterogeneity was evaluated by modeling RMST as a function of baseline predicted 6-month survival probability, including a treatment-by-risk interaction term in an overlap-weighted regression model.<br><br>Risk stratification: A crossover point r* was defined as the value of baseline predicted survival at which the estimated treatment effect reached a prespecified threshold of 30 days. Treatment effects were then estimated within strata defined by r < r* and r ≥ r* using stabilized IPTW-adjusted Kaplan–Meier curves and RMST. |
| Identifying assumptions | Randomization at treatment initiation ensures exchangeability, consistency, and positivity by design. Censoring is assumed to be non-informative. | We assume conditional exchangeability of treatment initiation given measured baseline covariates, consistency of treatment strategies, positivity within the region of clinical equipoise emphasized by overlap weighting, and non-informative censoring conditional on baseline covariates. |
| Data analysis plan | (1) Measure baseline covariates at or before time zero.<br><br>(2) Develop and calibrate a prognostic model to predict 6-month survival from baseline covariates, excluding treatment assignment.<br><br>(3) Estimate the effect of assignment to Strategy A versus Strategy B on 2-year RMST using an ITT analysis.<br><br>(4) Evaluate absolute treatment-effect heterogeneity by modeling RMST as a function of baseline predicted 6-month survival probability, including a treatment-by-risk interaction. Estimate treatment effect function TE(r) and corresponding confidence intervals.<br><br>(5) Summarize TE(r) using a prespecified crossover point r*, defined as the value of | (1) Measure baseline covariates at or before time zero.<br><br>(2) Develop and calibrate a prognostic model to predict 6-month survival from baseline covariates, excluding treatment assignment.<br><br>(3) Estimate the marginal effect of Strategy A versus Strategy B on 2-year RMST using stabilized IPTW.<br><br>(4) Evaluate absolute treatment-effect heterogeneity by modeling RMST as a function of baseline predicted 6-month survival probability, including a treatment-by-risk interaction, using overlap-weighted regression. Estimate treatment effect function TE(r) and corresponding confidence intervals. |

| | | |
|---|---|---|
| | baseline predicted survival at which the estimated treatment effect reaches 30 days of 2-year RMST. Report ITT effects within r < r* and r ≥ r* strata. | (5) Summarize TE(r) using a prespecified crossover point r*, defined as the value of baseline predicted survival at which the estimated treatment effect reaches 30 days of 2-year RMST. Estimate IPTW-adjusted survival and RMST within r < r* and r ≥ r* strata.<br><br>(6) Conduct sensitivity analyses to evaluate robustness to key analytic choices, including restriction to biomarker results available by treatment initiation, use of a ridge-penalized Cox prognostic model, a natural cubic spline treatment-effect specification, a 1-year RMST horizon, and alternative clinically meaningful RMST benefit thresholds of 14 and 60 days. |

**Table S4. Target trial emulation specifications for first-line pembrolizumab vs carboplatin plus gemcitabine in advanced urothelial carcinoma**

| TARGET component | Target trial specification | Emulation using observational data |
|---|---|---|
| Causal question | Primary causal question: among patients with advanced urothelial carcinoma (UC) initiating first-line therapy, how does the causal effect of carboplatin plus gemcitabine versus pembrolizumab on 2-year restricted mean survival time (RMST) vary with baseline predicted 6-month survival? | Same causal question addressed using the Flatiron Health Research Database. |
| Eligibility criteria | • Advanced UC<br>• Age ≥18<br>• Initiating first-line systemic therapy for advanced disease<br>• Eligibility is pragmatic and does not include strict laboratory or comorbidity exclusions | Eligibility was assessed at time zero (first-line treatment initiation) using diagnosis, staging, treatment, and biomarker data recorded in the Flatiron Health Research Database. Biomarker measurements reported within 30 days after treatment initiation were permitted to account for reporting delays. Patients initiating treatment too close to the data cutoff to permit adequate follow-up for the prespecified RMST horizon were excluded. |
| Treatment strategies | **Strategy A:** Initiate carboplatin plus gemcitabine<br><br>**Strategy B:** Initiate pembrolizumab | Treatment strategies were defined by the first-line systemic regimen recorded in the Flatiron Health Research Database.<br><br>**Strategy A:** Carboplatin plus gemcitabine<br><br>**Strategy B:** Pembrolizumab<br><br>In the chemotherapy arm, subsequent receipt of maintenance immunotherapy (e.g., avelumab) following completion of first-line chemotherapy was permitted and considered part of the chemotherapy treatment strategy, consistent with contemporary real-world practice. |
| Assignment procedures | Eligible patients are randomly assigned 1:1 to Strategy A versus Strategy B at time zero. Treatment initiation occurs immediately after assignment. | Treatment assignment was defined by the first-line regimen initiated at time zero, as recorded in the Flatiron Health Research Database. In the absence of randomization, analyses estimated a |

| | | |
|---|---|---|
| | | per-assignment (intention-to-treat [ITT] analogue) effect of treatment initiation without censoring for subsequent treatment discontinuation or switching, including receipt of maintenance immunotherapy. Confounding was addressed using propensity score methods to balance baseline covariates, with overlap weighting used for treatment-effect heterogeneity analyses and stabilized inverse probability of treatment weighting (IPTW) used for Kaplan–Meier and RMST estimation in the full cohort and within risk-defined strata. |
| Follow-up | Start: Time of randomization and treatment initiation (time zero).<br><br>End: Death, loss to follow-up, or administrative censoring at the planned end of the study period. | Start: Time of first-line systemic therapy initiation, defined as the earliest point at which eligibility and treatment assignment are simultaneously established.<br><br>End: Death or censoring at last recorded activity in the EHR. |
| Outcomes | Primary outcome: Overall survival from time zero.<br><br>Primary summary measure: RMST through 2 years from time zero. | Primary outcome: Overall survival from time zero.<br><br>Primary summary measure: RMST through 2 years from time zero. |
| Causal contrasts | Primary contrast: Difference in 2-year RMST under assignment to Strategy A versus Strategy B.<br><br>Heterogeneity analysis: Estimate the treatment-effect function TE(r) across baseline calibrated 6-month survival probability r using a treatment-by-risk interaction.<br><br>Risk stratification: Define a crossover point r* summarizing TE(r) at a clinically meaningful benefit threshold (30 days of 2-year RMST) and estimate treatment effects within $r < r^*$ and $r \geq r^*$ strata. | Primary contrast: Marginal difference in 2-year RMST between treatment strategies in the full cohort, estimated using stabilized IPTW.<br><br>Heterogeneity analysis: Treatment-effect heterogeneity was evaluated by modeling RMST as a function of baseline predicted 6-month survival probability, including a treatment-by-risk interaction term in an overlap-weighted regression model.<br><br>Risk stratification: A crossover point r* was defined as the value of baseline predicted survival at which the estimated treatment effect reached a |

| | | |
|---|---|---|
| | | prespecified threshold of 30 days. Treatment effects were then estimated within strata defined by $r < r^*$ and $r \geq r^*$ using stabilized IPTW-adjusted Kaplan–Meier curves and RMST. |
| Identifying assumptions | Randomization at treatment initiation ensures exchangeability, consistency, and positivity by design. Censoring is assumed to be non-informative. | We assume conditional exchangeability of treatment initiation given measured baseline covariates, consistency of treatment strategies, positivity within the region of clinical equipoise emphasized by overlap weighting, and non-informative censoring conditional on baseline covariates. |
| Data analysis plan | (1) Measure baseline covariates at or before time zero.<br><br>(2) Develop and calibrate a prognostic model to predict 6-month survival from baseline covariates, excluding treatment assignment.<br><br>(3) Estimate the effect of assignment to Strategy A versus Strategy B on 2-year RMST using an ITT analysis.<br><br>(4) Evaluate absolute treatment-effect heterogeneity by modeling RMST as a function of baseline predicted 6-month survival probability, including a treatment-by-risk interaction. Estimate treatment effect function TE(r) and corresponding confidence intervals.<br><br>(5) Summarize TE(r) using a prespecified crossover point $r^*$, defined as the value of baseline predicted survival at which the estimated treatment effect reaches 30 days of 2-year RMST. Report ITT effects within $r < r^*$ and $r \geq r^*$ strata. | (1) Measure baseline covariates at or before time zero.<br><br>(2) Develop and calibrate a prognostic model to predict 6-month survival from baseline covariates, excluding treatment assignment.<br><br>(3) Estimate the marginal effect of Strategy A versus Strategy B on 2-year RMST using stabilized IPTW.<br><br>(4) Evaluate absolute treatment-effect heterogeneity by modeling RMST as a function of baseline predicted 6-month survival probability, including a treatment-by-risk interaction, using overlap-weighted regression. Estimate treatment effect function TE(r) and corresponding confidence intervals.<br><br>(5) Summarize TE(r) using a prespecified crossover point $r^*$, defined as the value of baseline predicted survival at which the estimated treatment effect reaches 30 days of 2-year RMST. Estimate IPTW-adjusted survival and RMST within $r < r^*$ and $r \geq r^*$ strata.<br><br>(6) Conduct sensitivity analyses to evaluate robustness to key analytic choices, including restriction to |

| | |
|---|---|
| | biomarker results available by treatment initiation, use of a ridge-penalized Cox prognostic model, a natural cubic spline treatment-effect specification, a 1-year RMST horizon, and alternative clinically meaningful RMST benefit thresholds of 14 and 60 days. |

**Table S5. Target trial emulation specification for first-line pembrolizumab vs. chemotherapy in dMMR/MSI-H metastatic colorectal cancer**

| TARGET component | Target trial specification | Emulation using observational data |
|---|---|---|
| Causal question | Primary causal question: among patients with metastatic colorectal cancer (CRC) and dMMR/MSI-H initiating first-line therapy, how does the causal effect of pembrolizumab versus chemotherapy on 2-year restricted mean survival time (RMST) vary with baseline predicted 6-month survival? | Same causal question addressed using the Flatiron Health Research Database. |
| Eligibility criteria | • Metastatic CRC<br>• Age ≥18<br>• dMMR or MSI-H tumors<br>• Initiating first-line systemic therapy for advanced disease<br>• Eligibility is pragmatic and does not include strict laboratory or comorbidity exclusions | Eligibility was assessed at time zero (first-line treatment initiation) using diagnosis, staging, treatment, and biomarker data recorded in the Flatiron Health Research Database. Biomarker measurements reported within 30 days after treatment initiation were permitted to account for reporting delays. Patients initiating treatment too close to the data cutoff to permit adequate follow-up for the prespecified RMST horizon were excluded. |
| Treatment strategies | **Strategy A:** Initiate pembrolizumab monotherapy.<br><br>**Strategy B:** Initiate standard first-line fluoropyrimidine-based combination chemotherapy (FOLFOX, FOLFIRI, FOLFOXIRI, CAPEOX, or 5-fluorouracil/leucovorin), with or without biologic therapy (bevacizumab or cetuximab). | Treatment strategies were defined by the first-line systemic regimen initiated at time zero, as recorded in the Flatiron Health Research Database.<br><br>**Strategy A:** Pembrolizumab monotherapy.<br><br>**Strategy B:** Fluoropyrimidine-based combination chemotherapy (FOLFOX, FOLFIRI, FOLFOXIRI, CAPEOX, or 5-fluorouracil/leucovorin), with or without biologic therapy (bevacizumab or cetuximab).<br><br>Regimens including immune checkpoint inhibitors other than pembrolizumab or clinical trial drugs were excluded. |
| Assignment procedures | Eligible patients are randomly assigned 1:1 to Strategy A versus Strategy B at time zero. Treatment initiation occurs immediately after assignment. | Treatment assignment was defined by the first-line regimen initiated at time zero, as recorded in the Flatiron Health Research Database. In the absence of randomization, analyses estimated a per-assignment (intention-to-treat [ITT] analogue) effect of |

| | | |
|---|---|---|
| | | treatment initiation without censoring for subsequent treatment discontinuation or switching. Confounding was addressed using propensity score methods to balance baseline covariates, with overlap weighting used for treatment-effect heterogeneity analyses and stabilized inverse probability of treatment weighting (IPTW) used for Kaplan–Meier and RMST estimation in the full cohort and within risk-defined strata. |
| Follow-up | Start: Time of randomization and treatment initiation (time zero).<br><br>End: Death, loss to follow-up, or administrative censoring at the planned end of the study period. | Start: Time of first-line systemic therapy initiation, defined as the earliest point at which eligibility and treatment assignment are simultaneously established.<br><br>End: Death or censoring at last recorded activity in the EHR. |
| Outcomes | Primary outcome: Overall survival from time zero.<br><br>Primary summary measure: RMST through 2 years from time zero. | Primary outcome: Overall survival from time zero.<br><br>Primary summary measure: RMST through 2 years from time zero. |
| Causal contrasts | Primary contrast: Difference in 2-year RMST under assignment to Strategy A versus Strategy B.<br><br>Heterogeneity analysis: Estimate the treatment-effect function TE(r) across baseline calibrated 6-month survival probability r using a treatment-by-risk interaction.<br><br>Risk stratification: Define a crossover point r* summarizing TE(r) at a clinically meaningful benefit threshold (30 days of 2-year RMST) and estimate treatment effects within $r < r^*$ and $r \geq r^*$ strata. | Primary contrast: Marginal difference in 2-year RMST between treatment strategies in the full cohort, estimated using stabilized IPTW.<br><br>Heterogeneity analysis: Treatment-effect heterogeneity was evaluated by modeling RMST as a function of baseline predicted 6-month survival probability, including a treatment-by-risk interaction term in an overlap-weighted regression model.<br><br>Risk stratification: A crossover point r* was defined as the value of baseline predicted survival at which the estimated treatment effect reached a prespecified threshold of 30 days. Treatment effects were then estimated within strata defined by $r < r^*$ and $r \geq r^*$ using stabilized IPTW-adjusted Kaplan–Meier curves and RMST. |
| Identifying assumptions | Randomization at treatment initiation ensures exchangeability, consistency, and positivity by | We assume conditional exchangeability of treatment initiation given measured baseline covariates, consistency of treatment |

| | | |
|---|---|---|
| | design. Censoring is assumed to be non-informative. | strategies, positivity within the region of clinical equipoise emphasized by overlap weighting, and non-informative censoring conditional on baseline covariates. |
| Data analysis plan | (1) Measure baseline covariates at or before time zero.<br><br>(2) Develop and calibrate a prognostic model to predict 6-month survival from baseline covariates, excluding treatment assignment.<br><br>(3) Estimate the effect of assignment to Strategy A versus Strategy B on 2-year RMST using an ITT analysis.<br><br>(4) Evaluate absolute treatment-effect heterogeneity by modeling RMST as a function of baseline predicted 6-month survival probability, including a treatment-by-risk interaction. Estimate treatment effect function TE(r) and corresponding confidence intervals.<br><br>(5) Summarize TE(r) using a prespecified crossover point r*, defined as the value of baseline predicted survival at which the estimated treatment effect reaches 30 days of 2-year RMST. Report ITT effects within r < r* and r ≥ r* strata. | (1) Measure baseline covariates at or before time zero.<br><br>(2) Develop and calibrate a prognostic model to predict 6-month survival from baseline covariates, excluding treatment assignment.<br><br>(3) Estimate the marginal effect of Strategy A versus Strategy B on 2-year RMST using stabilized IPTW.<br><br>(4) Evaluate absolute treatment-effect heterogeneity by modeling RMST as a function of baseline predicted 6-month survival probability, including a treatment-by-risk interaction, using overlap-weighted regression. Estimate treatment effect function TE(r) and corresponding confidence intervals.<br><br>(5) Summarize TE(r) using a prespecified crossover point r*, defined as the value of baseline predicted survival at which the estimated treatment effect reaches 30 days of 2-year RMST. Estimate IPTW-adjusted survival and RMST within r < r* and r ≥ r* strata.<br><br>(6) Conduct sensitivity analyses to evaluate robustness to key analytic choices, including restriction to biomarker results available by treatment initiation, use of a ridge-penalized Cox prognostic model, prognostic model development restricted to chemotherapy-treated patients, a natural cubic spline treatment-effect specification, a 1-year RMST horizon, and alternative clinically meaningful RMST benefit thresholds of 14 and 60 days. |

**Table S6. Target trial emulation specifications for first-line nivolumab plus ipilimumab versus BRAF/MEK inhibitor combination therapy in BRAF-mutant advanced melanoma**

| TARGET component | Target trial specification | Emulation using observational data |
|---|---|---|
| Causal question | Primary causal question: among patients with advanced melanoma and BRAF-mutant disease initiating first-line therapy, how does the causal effect of ipilimumab plus nivolumab versus BRAF/MEK inhibitor combination therapy on 2-year restricted mean survival time (RMST) vary with baseline predicted 6-month survival? | Same causal question addressed using the Flatiron Health Research Database. |
| Eligibility criteria | • Advanced melanoma<br>• Age ≥18<br>• BRAF-mutant disease<br>• Initiating first-line systemic therapy for advanced disease<br>• No prior checkpoint inhibitor or BRAF-MEK–targeted therapy in the adjuvant or neoadjuvant setting<br>• Eligibility is pragmatic and does not include strict laboratory or comorbidity exclusions | Eligibility was assessed at time zero (first-line treatment initiation) using diagnosis, staging, treatment, and biomarker data recorded in the Flatiron Health Research Database. Biomarker measurements reported within 30 days after treatment initiation were permitted to account for reporting delays. Patients initiating treatment too close to the data cutoff to permit adequate follow-up for the prespecified RMST horizon were excluded. |
| Treatment strategies | **Strategy A:** Initiate nivolumab plus ipilimumab<br><br>**Strategy B:** Initiate BRAF-MEK inhibitor combinations (dabrafenib plus trametinib, encorafenib plus binimetinib, or vemurafenib plus cobimetinib) | Treatment strategies were defined by the first-line systemic regimen recorded in the Flatiron Health Research Database.<br><br>**Strategy A:** Nivolumab plus ipilimumab<br><br>**Strategy B:** BRAF-MEK inhibitor combinations (dabrafenib plus trametinib, encorafenib plus binimetinib, or vemurafenib plus cobimetinib)<br><br>Regimens including immune checkpoint inhibitors other than nivolumab or ipilimumab or clinical trial drugs were excluded. |
| Assignment procedures | Eligible patients are randomly assigned 1:1 to Strategy A versus Strategy B at time zero. | Treatment assignment was defined by the first-line regimen initiated at time |

| | | |
|---|---|---|
| | Treatment initiation occurs immediately after assignment. | zero, as recorded in the Flatiron Health Research Database. In the absence of randomization, analyses estimated a per-assignment (intention-to-treat [ITT] analogue) effect of treatment initiation without censoring for subsequent treatment discontinuation or switching. Confounding was addressed using propensity score methods to balance baseline covariates, with overlap weighting used for treatment-effect heterogeneity analyses and stabilized inverse probability of treatment weighting (IPTW) used for Kaplan–Meier and RMST estimation in the full cohort and within risk-defined strata. |
| Follow-up | Start: Time of randomization and treatment initiation (time zero).<br><br>End: Death, loss to follow-up, or administrative censoring at the planned end of the study period. | Start: Time of first-line systemic therapy initiation, defined as the earliest point at which eligibility and treatment assignment are simultaneously established.<br><br>End: Death or censoring at last recorded activity in the EHR. |
| Outcomes | Primary outcome: Overall survival from time zero.<br><br>Primary summary measure: RMST through 2 years from time zero. | Primary outcome: Overall survival from time zero.<br><br>Primary summary measure: RMST through 2 years from time zero. |
| Causal contrasts | Primary contrast: Difference in 2-year RMST under assignment to Strategy A versus Strategy B.<br><br>Heterogeneity analysis: Estimate the treatment-effect function TE(r) across baseline calibrated 6-month survival probability r using a treatment-by-risk interaction.<br><br>Risk stratification: Define a crossover point r* summarizing TE(r) at a clinically meaningful benefit threshold (30 days of 2-year RMST) and estimate treatment effects within r < r* and r≥ r* strata. | Primary contrast: Marginal difference in 2-year RMST between treatment strategies in the full cohort, estimated using stabilized IPTW.<br><br>Heterogeneity analysis: Treatment-effect heterogeneity was evaluated by modeling RMST as a function of baseline predicted 6-month survival probability, including a treatment-by-risk interaction term in an overlap-weighted regression model.<br><br>Risk stratification: A crossover point r* was defined as the value of baseline |

| | | |
|---|---|---|
| | | predicted survival at which the estimated treatment effect reached a prespecified threshold of 30 days. Treatment effects were then estimated within strata defined by r < r* and r ≥ r* using stabilized IPTW-adjusted Kaplan–Meier curves and RMST. |
| Identifying assumptions | Randomization at treatment initiation ensures exchangeability, consistency, and positivity by design. Censoring is assumed to be non-informative. | We assume conditional exchangeability of treatment initiation given measured baseline covariates, consistency of treatment strategies, positivity within the region of clinical equipoise emphasized by overlap weighting, and non-informative censoring conditional on baseline covariates. |
| Data analysis plan | (1) Measure baseline covariates at or before time zero.<br><br>(2) Develop and calibrate a prognostic model to predict 6-month survival from baseline covariates, excluding treatment assignment.<br><br>(3) Estimate the effect of assignment to Strategy A versus Strategy B on 2-year RMST using an ITT analysis.<br><br>(4) Evaluate absolute treatment-effect heterogeneity by modeling RMST as a function of baseline predicted 6-month survival probability, including a treatment-by-risk interaction. Estimate treatment effect function TE(r) and corresponding confidence intervals.<br><br>(5) Summarize TE(r) using a prespecified crossover point r*, defined as the value of baseline predicted survival at which the estimated treatment effect reaches 30 days of 2-year RMST. Report ITT effects within r < r* and r ≥ r* strata. | (1) Measure baseline covariates at or before time zero.<br><br>(2) Develop and calibrate a prognostic model to predict 6-month survival from baseline covariates, excluding treatment assignment.<br><br>(3) Estimate the marginal effect of Strategy A versus Strategy B on 2-year RMST using stabilized IPTW.<br><br>(4) Evaluate absolute treatment-effect heterogeneity by modeling RMST as a function of baseline predicted 6-month survival probability, including a treatment-by-risk interaction, using overlap-weighted regression. Estimate treatment effect function TE(r) and corresponding confidence intervals.<br><br>(5) Summarize TE(r) using a prespecified crossover point r*, defined as the value of baseline predicted survival at which the estimated treatment effect reaches 30 days of 2-year RMST. Estimate IPTW-adjusted survival and RMST |

| | |
|---|---|
| | within r < r* and r ≥ r* strata.<br><br>(6) Conduct sensitivity analyses to evaluate robustness to key analytic choices, including restriction to biomarker results available by treatment initiation, use of a ridge-penalized Cox prognostic model, a natural cubic spline treatment-effect specification, a 1-year RMST horizon, and alternative clinically meaningful RMST benefit thresholds of 14 and 60 days. |

**Table S7. Target trial emulation specifications in first-line nivolumab plus ipilimumab versus single-agent antiangiogenic therapy in metastatic renal cell carcinoma**

| TARGET component | Target trial specification | Emulation using observational data |
|---|---|---|
| Causal question | Primary causal question: among patients with clear cell or histology NOS metastatic renal cell carcinoma (RCC) initiating first-line therapy, how does the causal effect of ipilimumab plus nivolumab versus single-agent antiangiogenic therapy on 3-year restricted mean survival time (RMST) vary with baseline predicted 6-month survival? | Same causal question addressed using the Flatiron Health Research Database. |
| Eligibility criteria | • Metastatic RCC<br>• Clear cell histology or histology NOS<br>• Age ≥18<br>• Initiating first-line systemic therapy for metastatic disease<br>• Eligibility is pragmatic and does not include strict laboratory or comorbidity exclusions | Eligibility was assessed at time zero (first-line treatment initiation) using diagnosis, staging, treatment, and biomarker data recorded in the Flatiron Health Research Database. Biomarker measurements reported within 30 days after treatment initiation were permitted to account for reporting delays. Patients initiating treatment too close to the data cutoff to permit adequate follow-up for the prespecified RMST horizon were excluded. |
| Treatment strategies | **Strategy A:** Initiate nivolumab plus ipilimumab<br><br>**Strategy B:** Initiate single-agent VEGF-targeted antiangiogenic therapy (pazopanib or sunitinib). | Treatment strategies were defined by the first-line systemic regimen recorded in the Flatiron Health Research Database.<br><br>**Strategy A:** Nivolumab plus ipilimumab<br><br>**Strategy B:** Single-agent VEGF-targeted antiangiogenic therapy (pazopanib or sunitinib).<br><br>Regimens including immune checkpoint inhibitors other than nivolumab or ipilimumab, combination antiangiogenic regimens, or clinical trial drugs were excluded. |
| Assignment procedures | Eligible patients are randomly assigned 1:1 to Strategy A versus Strategy B at time zero. Treatment initiation occurs immediately after assignment. | Treatment assignment was defined by the first-line regimen initiated at time zero, as recorded in the Flatiron Health Research Database. In the absence of randomization, analyses estimated a per-assignment (intention-to-treat [ITT] analogue) effect of |

| | | |
|---|---|---|
| | | treatment initiation without censoring for subsequent treatment discontinuation or switching. Confounding was addressed using propensity score methods to balance baseline covariates, with overlap weighting used for treatment-effect heterogeneity analyses and stabilized inverse probability of treatment weighting (IPTW) used for Kaplan–Meier and RMST estimation in the full cohort and within risk-defined strata. |
| Follow-up | Start: Time of randomization and treatment initiation (time zero).<br><br>End: Death, loss to follow-up, or administrative censoring at the planned end of the study period. | Start: Time of first-line systemic therapy initiation, defined as the earliest point at which eligibility and treatment assignment are simultaneously established.<br><br>End: Death or censoring at last recorded activity in the EHR. |
| Outcomes | Primary outcome: Overall survival from time zero.<br><br>Primary summary measure: RMST through 3 years from time zero. | Primary outcome: Overall survival from time zero.<br><br>Primary summary measure: RMST through 3 years from time zero. |
| Causal contrasts | Primary contrast: Difference in 3-year RMST under assignment to Strategy A versus Strategy B.<br><br>Heterogeneity analysis: Estimate the treatment-effect function TE(r) across baseline calibrated 6-month survival probability r using a treatment-by-risk interaction.<br><br>Risk stratification: Define a crossover point r* summarizing TE(r) at a clinically meaningful benefit threshold (30 days of 3-year RMST) and estimate treatment effects within r < r* and r≥ r* strata. | Primary contrast: Marginal difference in 3-year RMST between treatment strategies in the full cohort, estimated using stabilized IPTW.<br><br>Heterogeneity analysis: Treatment-effect heterogeneity was evaluated by modeling RMST as a function of baseline predicted 6-month survival probability, including a treatment-by-risk interaction term in an overlap-weighted regression model.<br><br>Risk stratification: A crossover point r* was defined as the value of baseline predicted survival at which the estimated treatment effect reached a prespecified threshold of 30 days. Treatment effects were then estimated within strata defined by r < r* and r ≥ r* using stabilized IPTW-adjusted Kaplan–Meier curves and RMST. |
| Identifying assumptions | Randomization at treatment initiation ensures exchangeability, consistency, and | We assume conditional exchangeability of treatment initiation given measured baseline covariates, consistency of |

| | | |
|---|---|---|
| | positivity by design. Censoring is assumed to be non-informative. | treatment strategies, positivity within the region of clinical equipoise emphasized by overlap weighting, and non-informative censoring conditional on baseline covariates. |
| Data analysis plan | (1) Measure baseline covariates at or before time zero.<br><br>(2) Develop and calibrate a prognostic model to predict 6-month survival from baseline covariates, excluding treatment assignment.<br><br>(3) Estimate the effect of assignment to Strategy A versus Strategy B on 3-year RMST using an ITT analysis.<br><br>(4) Evaluate absolute treatment-effect heterogeneity by modeling RMST as a function of baseline predicted 6-month survival probability, including a treatment-by-risk interaction. Estimate treatment effect function TE(r) and corresponding confidence intervals.<br><br>(5) Summarize TE(r) using a prespecified crossover point r*, defined as the value of baseline predicted survival at which the estimated treatment effect reaches 30 days of 3-year RMST. Report ITT effects within $r < r^*$ and $r \geq r^*$ strata. | (1) Measure baseline covariates at or before time zero.<br><br>(2) Develop and calibrate a prognostic model to predict 6-month survival from baseline covariates, excluding treatment assignment.<br><br>(3) Estimate the marginal effect of Strategy A versus Strategy B on 3-year RMST using stabilized IPTW.<br><br>(4) Evaluate absolute treatment-effect heterogeneity by modeling RMST as a function of baseline predicted 6-month survival probability, including a treatment-by-risk interaction, using overlap-weighted regression. Estimate treatment effect function TE(r) and corresponding confidence intervals.<br><br>(5) Summarize TE(r) using a prespecified crossover point r*, defined as the value of baseline predicted survival at which the estimated treatment effect reaches 30 days of 3-year RMST. Estimate IPTW-adjusted survival and RMST within $r < r^*$ and $r \geq r^*$ strata. |

**Selection and specification of treatment comparisons**

Candidate treatment comparisons were identified among patients with advanced or metastatic solid tumors initiating first-line systemic therapy in the Flatiron Health Research Database. Comparisons were eligible when one treatment strategy used ICI therapy without chemotherapy or targeted therapy, including either single-agent or dual-ICI therapy. The alternative strategy was required to include a systemic therapy expected to provide more reliable early disease control, such as chemotherapy or targeted therapy, and could also include ICI. Comparisons in which both groups received chemotherapy or targeted therapy were excluded because early disease control from these agents in both groups could obscure the hypothesized excess early deaths with ICI alone.

We pragmatically required at least 300 patients in each treatment group to limit the inclusion of comparisons with sparse treatment arms. Comparisons were also required to have sufficient follow-up for estimation of RMST through the 2-year prespecified horizon. The candidate comparisons and final analytic designation are summarized in Table S8.

**Table S8. Evaluated first-line ICI comparisons and inclusion status**

| Comparison | Status |
|---|---|
| Recurrent/Metastatic HNSCC, CPS ≥1 or unknown PD-L1: pembrolizumab plus chemotherapy vs. pembrolizumab | Included |
| Advanced NSCLC, high PD-L1 (TPS ≥ 50%): pembrolizumab plus chemotherapy vs. pembrolizumab | Included |
| Advanced Urothelial: carboplatin plus gemcitabine vs pembrolizumab | Included |
| Metastatic Colorectal, dMMR/MSI-H: pembrolizumab vs. chemotherapy | Included |
| Advanced Melanoma, BRAF-mutant: ipilimumab plus nivolumab vs. BRAF-MEK inhibitors | Included |
| Metastatic RCC, clear cell: ipilimumab plus nivolumab vs. single-agent antiangiogenic therapy | Included |
| Advanced NSCLC, low PD-L1 (TPS 1-49%): ipilimumab plus nivolumab vs. pembrolizumab plus chemotherapy | Excluded |
| Metastatic RCC, clear cell: ipilimumab plus nivolumab vs. ICI plus antiangiogenic therapy | Excluded |
| Metastatic BC, triple negative: pembrolizumab vs. chemotherapy | Excluded |
| Metastatic BC, triple negative: pembrolizumab plus chemotherapy vs. pembrolizumab | Excluded |
| Metastatic Gastric/GEJ, CPS ≥1, HER2 negative: pembrolizumab plus chemotherapy vs. pembrolizumab | Excluded |
| Metastatic HCC: nivolumab vs. sorafenib | Excluded |

Five comparisons were included in the primary analysis. Metastatic renal cell carcinoma (RCC) was analyzed separately as a concordance cohort because CheckMate 214 provided an established, prognosis-associated pattern of treatment-effect heterogeneity against which the framework could be evaluated. Eligibility criteria, treatment strategies, follow-up, causal contrasts, identifying assumptions, and analysis plans are provided in Tables S2-S7. Figure S1 shows cohort derivation and treatment assignment.

**Figure S1. CONSORT-style flow diagrams for study cohorts**

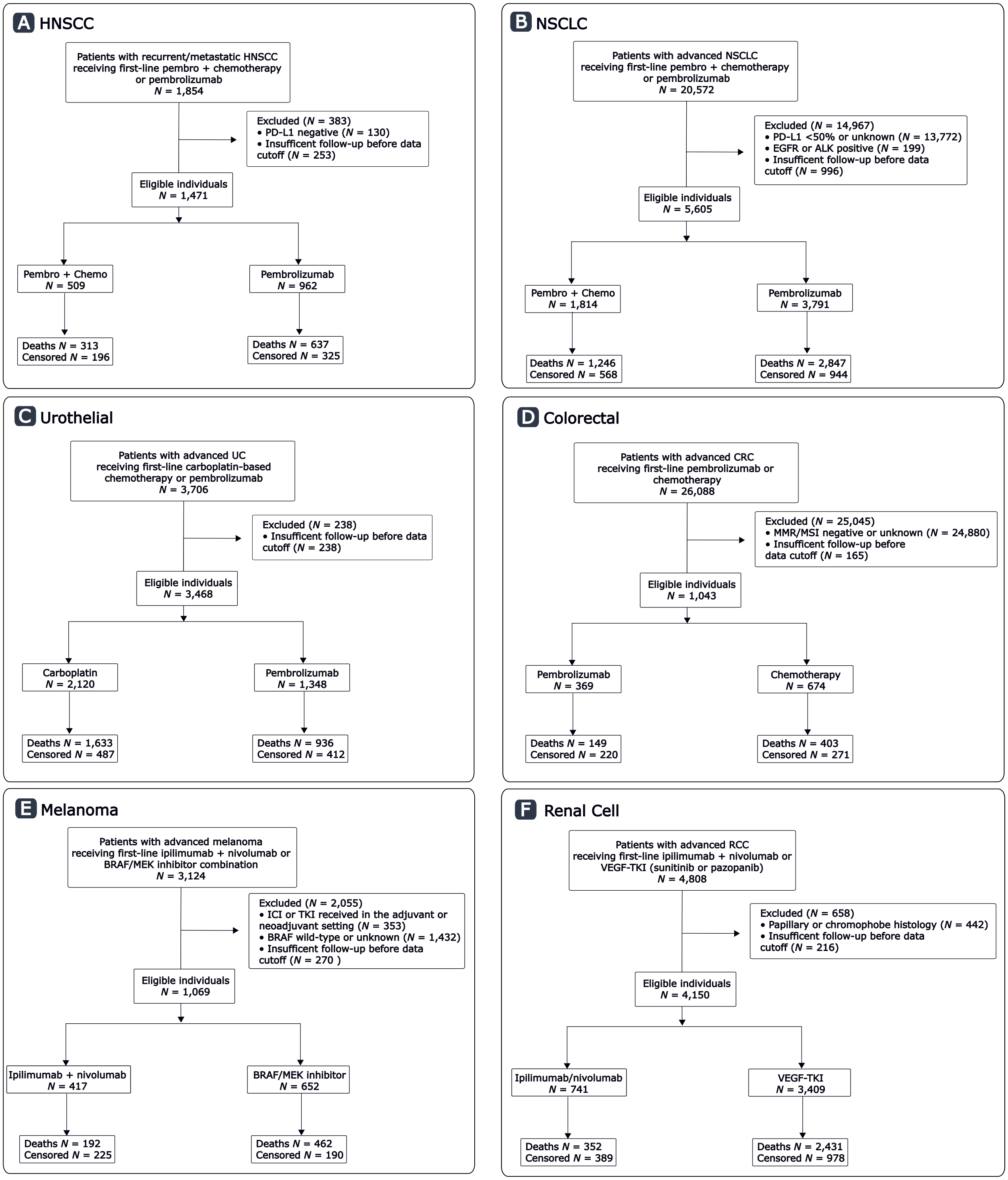

Cohort-specific treatment-initiation windows and data-lock dates are shown in Table S9. Follow-up availability was considered at the cohort level; individual patients were not required to have potential follow-up covering the full RMST horizon. The secondary RCC concordance analysis used an older data extract with an October 2021 data lock and included treatment initiation through 2020. Consequently, patients initiating treatment later in this window had potential follow-up shorter than the 3-year RMST horizon, and estimation at later time points relied on patients with longer available follow-up.

**Table S9. Observation windows and data cutoff dates by cohort**

| Cohort | First-line treatment window | Data lock |
|---|---|---|
| HNSCC | 2015-2022 | Oct 2023 |
| NSCLC | 2015-2023 | Nov 2025 |
| Urothelial | 2011-2022 | Oct 2023 |
| Colorectal | 2013-2023 | Nov 2025 |
| Melanoma | 2013-2021 | Oct 2023 |
| RCC | 2011-2020 | Oct 2021 |

**Feature construction and missing data handling**

The index date was defined as initiation of first-line therapy for advanced or metastatic disease. Follow-up began at the index date and continued until death. Patients without a recorded death date were right-censored at their last documented activity in the EHR.

Baseline clinical features were measured within the 90 days preceding and including the index date, with the value closest to the index date selected when multiple measurements were available. Biomarker variables were handled separately because the Flatiron Health Research Database records both specimen collection date and result date. Consistent with Flatiron guidance, biomarker timing was defined using the result date, which reflects when the finalized result became available in the EHR. Because result availability frequently lagged specimen collection by several weeks, with a typical delay of approximately 20–30 days in the study data, biomarker results reported within 30 days after the index date were permitted. This window was intended to capture delayed reporting of specimens collected before or near treatment initiation and reduce misclassification of cohort eligibility, rather than incorporate biological changes arising after treatment. For example, it allowed identification of patients with EGFR-positive NSCLC whose results were reported after treatment began.

Categorical variables were one-hot encoded, with missingness represented as a separate category. ECOG performance status and stage were retained as ordinal variables and imputed using the modal category. Continuous variables were median-imputed, and a corresponding missingness indicator was included for each imputed variable. The final feature counts were 115 for HNSCC, 122 for NSCLC, 118 for urothelial, 115 for colorectal, 126 for melanoma, and 117 for RCC.

**Prognostic model training and discrimination**

The goal of the prognostic model was to estimate the probability of 6-month survival from the start of first-line therapy for advanced disease. We trained a gradient-boosted survival model optimizing the Cox proportional hazards partial likelihood, implemented in scikit-survival (Python 3.13). Model selection was based on prior benchmarking in Flatiron EHR data demonstrating superior prognostic discrimination compared with alternative ML modeling approaches [2].

Each cancer dataset was randomly divided into training (80%) and test (20%) subsets, stratified by death status to preserve event balance. Hyperparameters were optimized via 3-fold cross-validation on the training set to maximize the time-dependent area under the receiver operating characteristic curve (AUC) at 6 months, calculated from the model's predicted risk scores, using a hyperparameter search space as shown in table below. All other hyperparameters were set to their default values in the scikit-survival implementation.

**Table S10. Hyperparameter search space for gradient-boosted survival models**

| Hyperparameters | Candidate Values |
|---|---|
| Number of estimators | HNSCC, Urothelial, and Melanoma: 300, 500; Colorectal and RCC: 300, 500, 700; NSCLC: 400, 700 |
| Learning rate | 0.05, 0.10 |
| Maximum tree depth | 2, 3, 4 |
| Subsample ratio | 0.75, 0.85 |
| Feature sampling ratio | 0.70, 0.90 |

Model performance was then evaluated on the held-out test set using the same metric. Ninety-five percent confidence intervals (CIs) for AUC were obtained via 1,000 bootstrap iterations. After model tuning, out-of-sample 6-month survival probabilities were generated for all patients in the full dataset using cross-validation, with 5 folds for HNSCC, urothelial carcinoma, and melanoma and 3 folds for NSCLC and colorectal cancer. In each iteration, the model was trained on all but one fold and applied to the held-out fold, ensuring that each patient's survival prediction was generated from a model that had not been trained on that patient.

**Generation and calibration of out-of-sample predictions**

Predicted 6-month survival probabilities were calibrated using isotonic regression with cross-validation and applied to the study cohort of interest. For each fold, jackknife pseudo-observations for 6-month survival were computed from the training folds, and isotonic regression was used to map uncalibrated model predictions to these pseudo-observations. The fitted calibration model was then applied to the held-out fold to generate calibrated predictions. This process was repeated until all patients had calibrated, out-of-sample predictions.

Calibration performance was evaluated using the mean Brier score across test folds and by plotting predicted versus observed 6-month survival. Calibration slope and intercept were also reported, with values near 1 and 0, respectively, indicating good agreement.

**Estimation of the continuous treatment-effect function**

Jackknife pseudo-observations for 2-year RMST were then computed within the study cohort. Heterogeneity of absolute treatment benefit was evaluated using these pseudo-observations in an overlap-weighted least-squares regression model of the form:

$$RMST_{2yr,i} = \beta_0 + \beta_1 T_i + \beta_2 r_i + \beta_3 (T_i \; x \; r_i) + \varepsilon_i$$

where $i$ indexes individual patients, $T_i$ denotes treatment assignment, $r_i$ is the calibrated predicted probability of 6-month survival, and $\beta_3$ represents effect modification by baseline predicted survival.

The corresponding treatment-effect function was defined as:

$$TE(r) = \beta_1 + \beta_3 r$$

representing the expected difference in 2-year RMST between treatment arms at a given value of $r$. Predicted absolute treatment effects were evaluated across the observed continuum of calibrated 6-month survival probabilities using $\mathrm{TE}(r)$. Standard errors and 95% CIs for $\mathrm{TE}(r)$ were estimated using the delta method. Treatment-effect heterogeneity was assessed using the treatment-by-risk interaction term ($\beta_3$) in the overlap-weighted regression model with a two-sided Wald test (α = 0.05) using robust standard errors.

**Definition of the crossover point**

The primary RMST threshold, denoted δ, was 30 days. The crossover point ($r^*$) was the value of predicted 6-month survival at which the fitted treatment-effect function reached δ:

$$TE(r^*) = \delta, \text{ therefore } r^* = \frac{(\delta - \beta_1)}{\beta_3}.$$

A crossover point was reported only when the fitted treatment-effect function reached the threshold within the empirical range of predicted survival used for estimation.

##### Propensity score estimation and weighting

Propensity scores were estimated separately within each analytic cohort using L2-penalized logistic regression with the lbfgs solver, balanced class weights, and a maximum of 1,000 iterations. Propensity scores were constrained to [0.01, 0.99] before weight construction to limit numerical instability from predictions at the boundaries.

Three related analyses used the propensity score differently. Stabilized inverse probability of treatment weighting (IPTW) estimated marginal survival curves and RMST differences in the full cohort. Overlap weighting was used in the continuous treatment-effect heterogeneity model. For cohorts with an identified crossover point, stabilized IPTW was re-estimated within each risk stratum to estimate stratum-specific marginal survival and RMST.

The same cohort-specific covariate specification was used to construct overlap weights and stabilized IPTW. Covariates are listed in Table S11.

**Table S11. Cohort-specific propensity score variables**

| Cohort | Covariates |
|---|---|
| HNSCC | HPV status; PD-L1 status; age; sex; days from initial diagnosis to advanced disease; stage group; ECOG performance status and missingness; socioeconomic status and missingness; Medicaid insurance; treatment before 2020; treatment within 30 days of advanced diagnosis; prior surgery; prior radiation; local recurrence; distant recurrence; smoking history; percent weight change; albumin; total bilirubin; creatinine; van Walraven comorbidity score. |
| NSCLC | Histology; smoking history; age; sex; days from initial diagnosis to advanced disease; stage group; ECOG performance status and missingness; socioeconomic status and missingness; Medicaid insurance; treatment within 30 days of advanced diagnosis; percent weight change; hemoglobin; albumin; total bilirubin; creatinine; van Walraven comorbidity score. |
| Urothelial | Smoking history; PD-L1 status; age; sex; days from initial diagnosis to advanced disease; stage group and missingness; ECOG performance status and missingness; socioeconomic status and missingness; Medicaid insurance; treatment before 2020; treatment within 30 days of advanced diagnosis; prior surgery; lower-tract primary site; baseline weight; percent weight change; albumin; total bilirubin; creatinine; van Walraven comorbidity score. |
| Colorectal | Primary colorectal site; age; sex; days from initial diagnosis to metastatic disease; stage group; ECOG performance status and missingness; socioeconomic status and missingness; Medicaid insurance; treatment within 30 days of metastatic diagnosis; percent weight change; hemoglobin; albumin; alkaline phosphatase; carcinoembryonic antigen; van Walraven comorbidity score. |
| Melanoma | Age; sex; days from advanced diagnosis to treatment; stage group and missingness; ECOG performance status and missingness; socioeconomic status and missingness; Medicaid insurance; treatment before 2020; brain metastasis; liver metastasis; baseline weight; percent weight change; albumin; total bilirubin; lactate dehydrogenase; creatinine; van Walraven comorbidity score. |
| RCC | Stage group; age; days from initial diagnosis to metastatic disease; ECOG performance status and missingness; nephrectomy; smoking history; commercial insurance; Medicaid insurance; treatment within 1 year of metastatic diagnosis; baseline weight; percent weight change; albumin; creatinine; calcium; neutrophil count; platelet count; hemoglobin; van Walraven comorbidity score. |

##### RCT benchmarking

We compared full-cohort treatment effects with those reported in analogous RCTs: KEYNOTE-048 for HNSCC [3], PERSEE for NSCLC [4], KEYNOTE-361 for urothelial carcinoma [5], KEYNOTE-177 for colorectal cancer [6], and DREAMseq for melanoma [7]. Benchmarking used primary or coprimary overall survival endpoints, except for PERSEE, in which overall survival was a secondary endpoint. We used the treatment-effect measures reported in the corresponding RCTs: 2-year overall survival RMST differences for NSCLC, overall survival HRs for urothelial and colorectal cancer, and absolute differences in 2-year overall survival for melanoma. HRs were estimated using stabilized IPTW-weighted Cox proportional hazards models with robust standard errors. Concordance was assessed using three criteria adapted from the RCT-DUPLICATE framework: (1) significance agreement, defined as concordance in statistical significance and direction of effect; (2) 95% CI agreement, defined as the emulated estimate falling within the RCT's 95% CI; and (3) standardized-difference agreement, defined as an absolute standardized difference below 1.96 between the emulated and RCT estimates [8].

Because KEYNOTE-048 compared pembrolizumab monotherapy and pembrolizumab plus chemotherapy each with cetuximab plus chemotherapy, and did not report a direct comparison between the two pembrolizumab-containing arms, the HNSCC cohort was not included in formal concordance testing. Instead, we descriptively compared median overall survival in each treatment arm of the HNSCC cohort with the corresponding KEYNOTE-048 arms in the PD-L1 CPS ≥1 population.

**RCC concordance analysis**

We applied the analytic framework to metastatic RCC, comparing ipilimumab plus nivolumab with single-agent VEGF-targeted therapy to assess whether our approach could recover known treatment-effect heterogeneity. In CheckMate 214, ipilimumab plus nivolumab produced greater survival benefit among patients with International Metastatic Renal Cell Carcinoma Database Consortium (IMDC) defined intermediate- or poor-risk disease than among those with favorable-risk disease. We included clear-cell histology and histology recorded as not otherwise specified to retain sufficient numbers for analysis. Because overall survival remained immature at approximately 2 years of follow-up in CheckMate 214, a 3-year RMST horizon was used.

**Sensitivity analyses**

For each sensitivity analysis, we compared the treatment-effect difference between the 10th and 90th percentiles of baseline predicted survival, the treatment-by-risk interaction *P* value, and the predicted survival probability at which the estimated treatment effect crossed the 30-day RMST threshold ($r^*$), when a crossing occurred within the observed risk distribution.

*Biomarker result availability at treatment initiation*

To assess sensitivity to the biomarker timing window, we repeated the analysis using only biomarker results available on or before the index date, rather than allowing results reported through day 30. We reconstructed the biomarker-defined cohorts under this restriction and repeated prognostic modeling and downstream treatment-effect estimation. We also compared cohort sizes with those in the primary analysis.

*Alternative prognostic model*

To assess sensitivity to prognostic model choice, we replaced the gradient-boosted survival model with a ridge-penalized Cox proportional hazards model trained on the same covariates. The penalty strength was selected by 3-fold cross-validation to optimize 6-month AUC, consistent with the primary model's tuning procedure. We calibrated the resulting predictions and repeated the downstream treatment-effect analysis, also comparing model discrimination.

*Alternative prognostic model-development population*

To assess sensitivity to the prognostic model-development population, we retrained the gradient-boosted model using only chemotherapy-treated patients in two cohorts, HNSCC and colorectal cancer, rather than all first-line patients. We calibrated the resulting predictions and repeated the downstream treatment-effect analysis using the same measures. This analysis assessed whether including patients who received different treatment strategies during model development materially affected the estimated treatment-effect patterns.

*Alternative functional form*

To assess sensitivity to the assumed functional form, we modeled calibrated 6-month survival probability using a natural cubic spline with 3 degrees of freedom, allowing both the main effect of predicted survival and its interaction with treatment to vary flexibly. In place of the single treatment-by-risk interaction *P* value from the linear model, we used a joint Wald test of the treatment-by-spline terms to assess treatment-effect heterogeneity.

*Alternative RMST horizon*

We repeated the primary analysis using a 1-year RMST horizon in place of the prespecified 2-year horizon. We retained the 30-day threshold when estimating ($r^*$) to assess whether the direction of treatment-effect heterogeneity and the threshold crossing were preserved at the shorter horizon.

*Alternative clinically meaningful benefit thresholds*

To examine the influence of the clinically meaningful benefit threshold, we repeated the crossover analysis using thresholds of 14 and 60 days of 2-year RMST rather than the prespecified 30-day threshold.

*Analysis restricted to documented PD-L1 CPS ≥1 in HNSCC*

Finally, to assess whether the HNSCC treatment-by-risk pattern was also evident among patients with documented PD-L1 expression, we restricted the HNSCC cohort to patients with CPS ≥1 and repeated the overlap-weighted treatment-effect heterogeneity analysis using the calibrated prognostic scores. We compared the 10th–90th percentile treatment-effect contrast, interaction *P* value, and estimated 30-day RMST benefit threshold with those from the full HNSCC cohort.

**Exploratory Analyses**

*Subsequent therapies and censoring at ICI initiation*

We summarized treatment patterns following first-line therapy. In cohorts with frequent subsequent ICI use in the chemotherapy arms, we repeated the treatment-effect heterogeneity analyses after censoring chemotherapy-treated patients at initiation of second-line ICI therapy. Patients initially receiving ICI were followed regardless of subsequent treatment.

**Trial protocol registration**

This observational target trial emulation was not prospectively registered. The target trial protocols were publicly deposited after completion of the analyses at Zenodo (doi:10.5281/zenodo.23113983).

#### Supplementary Results

##### Prognostic model performance and calibration

Cancer-specific prognostic models were developed in populations that included all patients initiating first-line systemic therapy and were therefore substantially larger than the corresponding analytic treatment cohorts (Table S12).

**Table S12. Baseline characteristics of prognostic model populations across cancers**

| Cancer | Total *N* | Age, median (IQR) | Male, *N* (%) | ECOG ≥2, *N* (%) | Median follow-up (months) | Deaths, *N* (%) |
|---|---|---|---|---|---|---|
| HNSCC | 8,718 | 65 (58-73) | 6,718 (77.1) | 988 (16.5) | 41.3 | 6,187 (71.0) |
| NSCLC | 82,739 | 69 (62-76) | 42,363 (51.2) | 11,517 (21.0) | 56.5 | 61,287 (74.1) |
| Urothelial | 9,357 | 73 (66-79) | 6,863 (73.3) | 1,269 (20.3) | 42.4 | 6,415 (68.6) |
| Colorectal | 37,381 | 64 (54-73) | 20,895 (55.9) | 3,235 (13.3) | 53.7 | 25,250 (67.5) |
| Melanoma | 9,030 | 68 (58-76) | 6,052 (67.0) | 777 (13.7) | 41.1 | 4,724 (52.3) |
| RCC | 8,291 | 67 (59-74) | 5,841 (70.4) | 885 (19.0) | 42.6 | 5,085 (61.3) |

In held-out test data, discrimination for 6-month survival was consistent across cancer types, with AUCs ranging from 0.751 in HNSCC to 0.798 in melanoma (Table S13 and Figure S2). Training-set AUCs were modestly higher than test-set estimates in all cohorts except NSCLC, without evidence of substantial overfitting. Key predictors included performance status, laboratory markers, and measures of disease burden (Figure S2).

**Table S13. Prognostic model discrimination in training and held-out test data across cancers**

| Cancer | 6-Month AUC on Training Data (95% CI) | 6-Month AUC on Test Data (95% CI) |
|---|---|---|
| HNSCC | 0.803 (0.791-0.816) | 0.751 (0.724-0.777) |
| NSCLC | 0.773 (0.770-0.776) | 0.773 (0.769-0.776) |
| Urothelial | 0.838 (0.828-0.848) | 0.796 (0.775-0.818) |
| Colorectal | 0.813 (0.806-0.820) | 0.791 (0.776-0.805) |
| Melanoma | 0.860 (0.849-0.871) | 0.798 (0.773-0.823) |
| RCC | 0.820 (0.808-0.833) | 0.795 (0.768-0.820) |

Following isotonic calibration, predicted 6-month survival closely agreed with observed survival across cancer types (Figure S2). Predicted survival probabilities also spanned a broad range within each analytic cohort, approximately 0.3 to 0.9, supporting evaluation of treatment effects across clinically meaningful variation in baseline prognosis.

**Figure S2. Prognostic model discrimination and calibration**

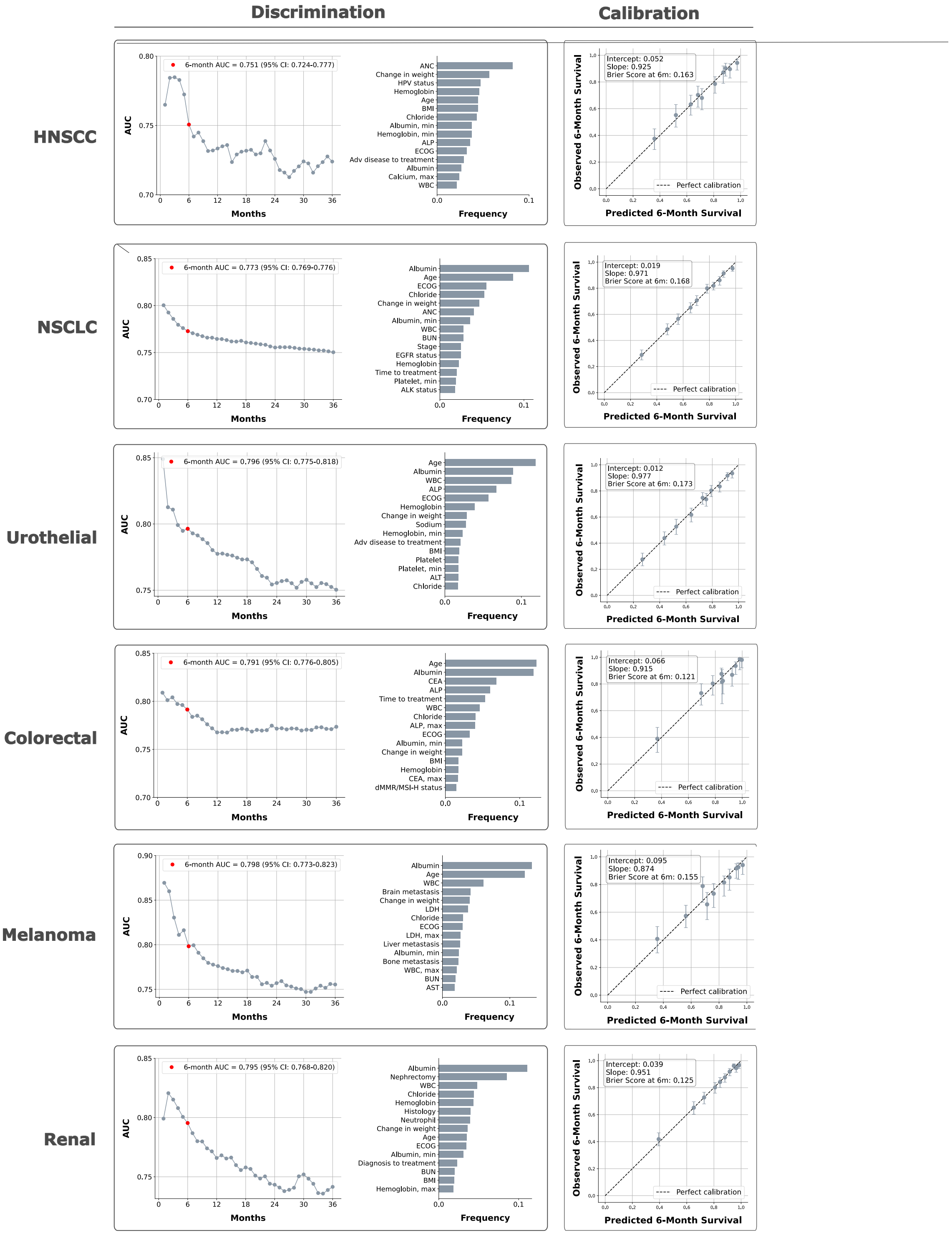

**Left column**: Time-dependent AUC estimated on the held-out test set, with the x-axis reflecting time from first-line systemic therapy initiation, alongside the 15 most important features in the gradient-boosted survival model ranked by frequency of use across trees. The prognostic model was trained on all patients with advanced disease receiving first-line systemic therapy within each cancer type, regardless of regimen. **Right column**: Calibration of predicted vs. observed 6-month survival after isotonic regression, evaluated specifically among patients in the analytic cohort of interest. Points represent the mean predicted and observed 6-month survival probability within each decile of predicted risk.

##### Weighting diagnostics and covariate balance

Arm-specific effective sample sizes after overlap weighting and stabilized IPTW are summarized in Table S14. Effective sample sizes remained above 300 in all treatment arms under both weighting approaches, with greater reductions generally observed under stabilized IPTW.

**Table S14. Arm-specific effective sample sizes (ESS) after weighting**

| Cohort | Arm | Raw *N* | Overlap ESS | IPTW ESS |
|---|---|---|---|---|
| HNSCC | Chemotherapy + Pembrolizumab | 509 | 425 | 403 |
| | Pembrolizumab | 962 | 815 | 727 |
| NSCLC | Chemotherapy + Pembrolizumab | 1,814 | 1,561 | 1,497 |
| | Pembrolizumab | 3,791 | 3,209 | 2,985 |
| Urothelial | Carboplatin/Gemcitabine | 2,120 | 1,724 | 1,269 |
| | Pembrolizumab | 1,348 | 1,094 | 1,072 |
| Colorectal | Pembrolizumab | 369 | 341 | 326 |
| | Chemotherapy | 674 | 628 | 629 |
| Melanoma | Ipilimumab/Nivolumab | 417 | 349 | 346 |
| | BRAF/MEK | 652 | 560 | 482 |

Propensity score distributions demonstrated substantial overlap between treatment groups across cohorts, and covariate balance was well achieved after weighting, with standardized mean differences typically <0.1 for variables using both IPTW and overlap weighting (Figure S3). Similar findings were observed within the high-risk strata of the risk-dependent cohorts, where propensity score overlap remained adequate and all standardized mean differences were <0.1 following stabilized IPTW (Figure S4). Missingness  of key baseline variables used in the propensity score models is summarized in Table S15.

**Table S15. Percent missingness of key baseline variables in study case cohorts**

| Cohort | ECOG | Hgb | WBC | Plt | Creatinine | T bili | ALP | AST | ALT | Albumin |
|---|---|---|---|---|---|---|---|---|---|---|
| HNSCC | 24.4 | 5.8 | 5.8 | 5.9 | 14.3 | 15.5 | 14.9 | 15.2 | 15.4 | 15.8 |
| NSCLC | 25.0 | 6.9 | 6.9 | 6.9 | 11.2 | 13.7 | 13.5 | 13.5 | 14.0 | 13.7 |
| Urothelial | 31.6 | 5.9 | 5.9 | 5.9 | 11.9 | 14.6 | 14.0 | 14.1 | 14.6 | 15.4 |
| Colorectal | 28.2 | 6.5 | 6.5 | 6.7 | 10.9 | 12.7 | 12.3 | 12.3 | 12.5 | 3.7 |
| Melanoma | 32.7 | 16.9 | 16.9 | 16.9 | 21.4 | 21.7 | 21.1 | 21.0 | 21.2 | 24.4 |
| RCC | 47.2 | 22.9 | 23.0 | 23.2 | 31.5 | 33.5 | 32.3 | 32.4 | 32.8 | 34.4 |

*Hgb = Hemoglobin; Plt = Platelet; T bili = Total bilirubin*

#### Figure S3. Propensity score overlap and covariate balance diagnostics across study cohorts

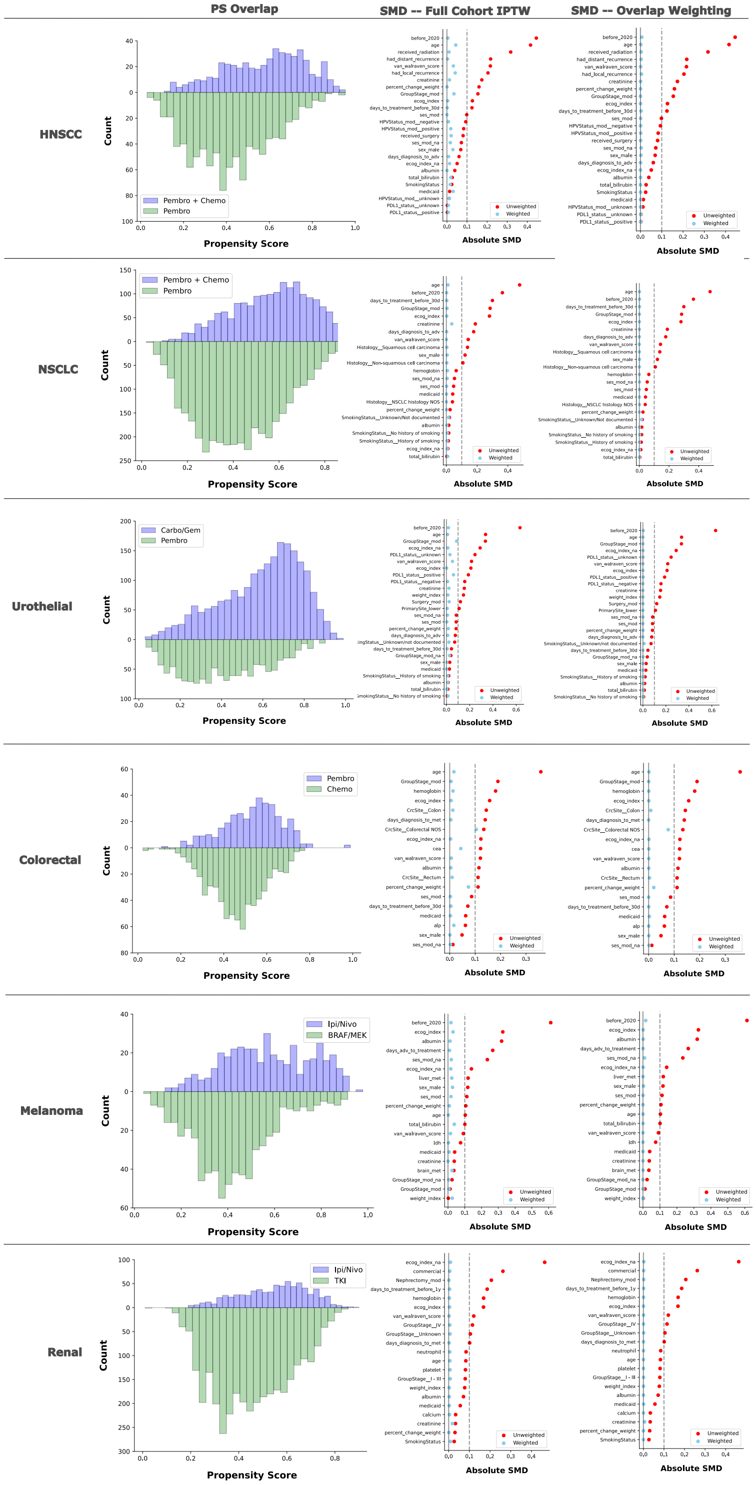

Propensity score (PS) distributions and standardized mean differences (SMDs) are shown for each cohort. The PS overlap column displays the distribution of estimated propensity scores by treatment arm before weighting. The full-cohort IPTW column displays covariate balance before and after stabilized inverse probability of treatment weighting (IPTW), used for Kaplan–Meier and restricted mean survival time estimation. The overlap weighting column displays covariate balance before and after overlap weighting, used for treatment-effect heterogeneity modeling. The same propensity score model underlies both weighting approaches. Dashed vertical lines indicate an absolute SMD of 0.10, a commonly used threshold for adequate covariate balance.

#### Figure S4. Propensity score overlap and covariate balance diagnostics within high-risk strata

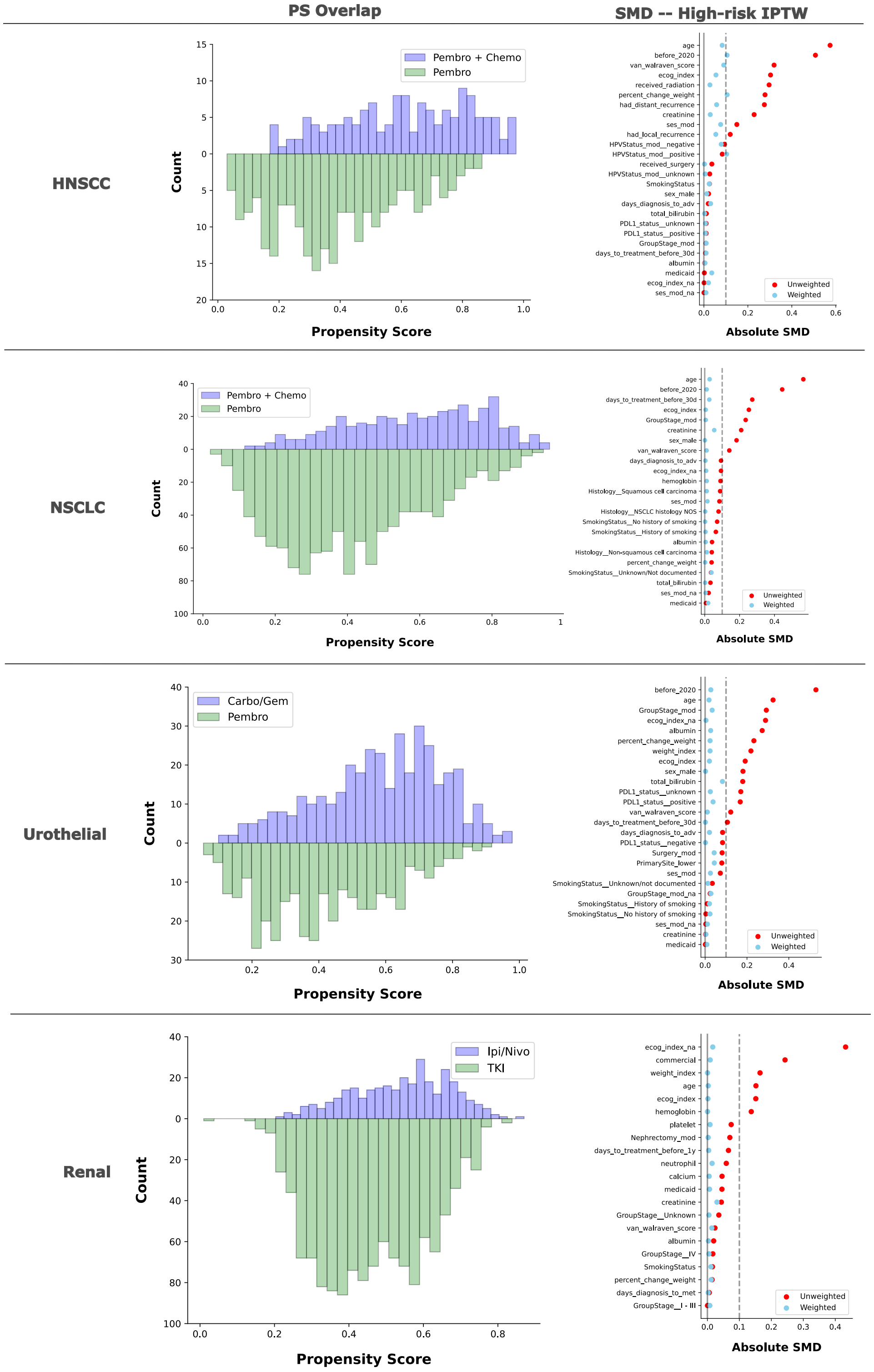

Propensity score (PS) distributions and standardized mean differences (SMDs) are shown for the high-risk stratum of each risk dependent cohort (HNSCC, NSCLC, urothelial carcinoma, and RCC). High-risk strata were defined using the cohort-specific crossover threshold (r*). The PS overlap column displays the distribution of estimated propensity scores by treatment arm before weighting. The high-risk IPTW column displays covariate balance before and after stabilized inverse probability of treatment weighting (IPTW). Dashed vertical lines indicate an absolute SMD of 0.10, a commonly used threshold for adequate covariate balance.

##### Treatment-effect heterogeneity by baseline predicted survival

Cohort-specific treatment-effect analyses for advanced NSCLC with PD-L1 TPS ≥50%, urothelial carcinoma, and BRAF-mutant melanoma are shown in Figures S5–S7. Baseline characteristics by risk stratum for the HNSCC, NSCLC, and urothelial carcinoma cohorts are shown in Tables S16–S18.

**Figure S5. Heterogeneity of chemotherapy benefit by baseline predicted survival in advanced NSCLC with PD-L1 TPS ≥50%**

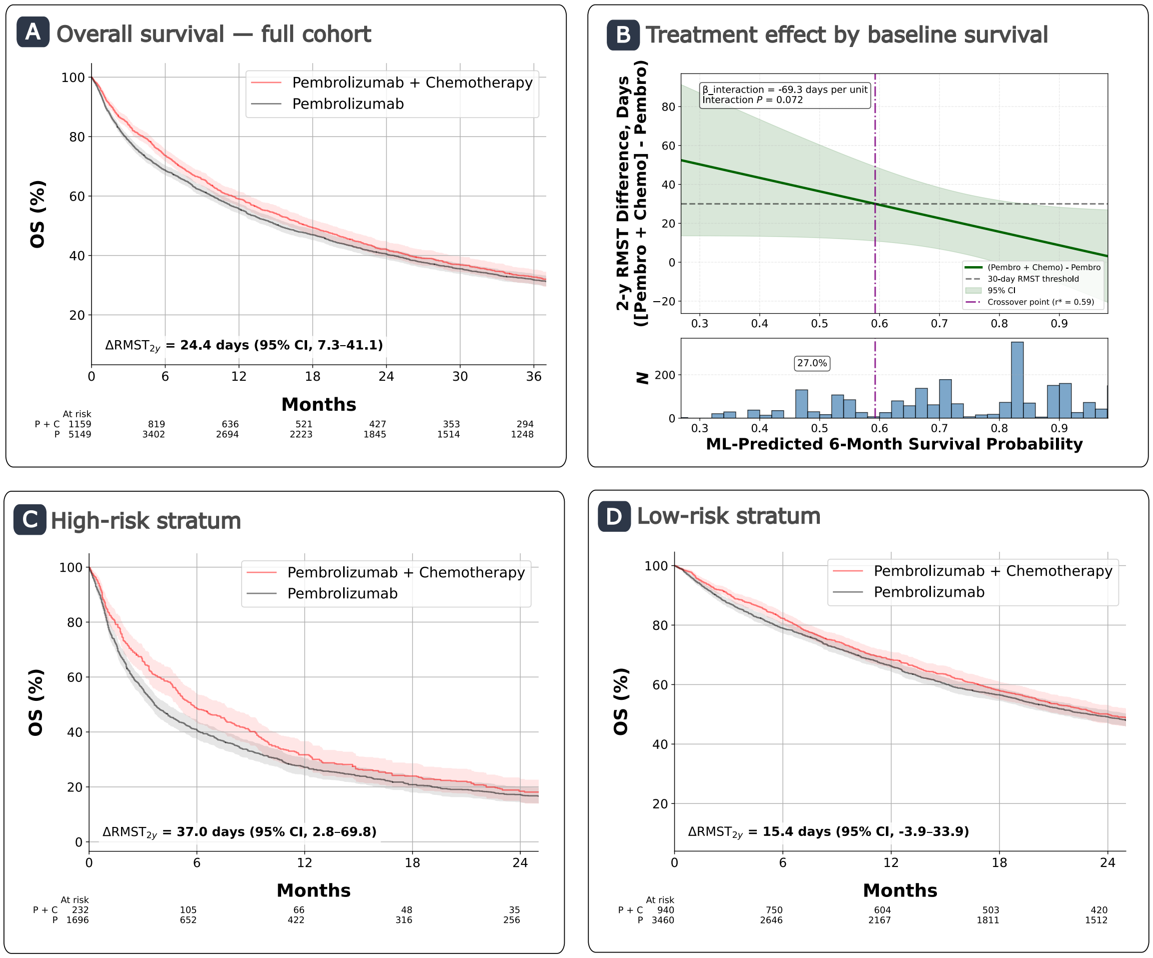

**Panel A**: IPTW-adjusted overall survival in the full cohort, showing an estimated 2-year RMST difference of 24.4 days between pembrolizumab plus chemotherapy and pembrolizumab monotherapy (95% CI: 7.3 to 41.1).

**Panel B**: Estimated treatment-effect function demonstrating increased benefit of combination therapy as baseline predicted survival worsened, with a borderline treatment-by-risk interaction ($\beta_{interaction}$ = −6.9 days per 0.1 increase in predicted 6-month survival, $P$ = .07). The crossover point (r*), defined as the predicted survival probability at which combination therapy confers a ≥ 30-day improvement in 2-year RMST, occurred at r = 0.59.

**Panels C and D**: IPTW-adjusted Kaplan–Meier curves stratified by r*. Among higher-risk patients (r < 0.59), combination therapy improved survival ($\Delta RMST_{2y}$ = 37.0 days, 95% CI: 2.8 to 69.8). Among lower-risk patients (r ≥ 0.59), the benefit was smaller and not statistically significant ($\Delta RMST_{2y}$ = 15.4 days, 95% CI: -3.9 to 33.9). Numbers at risk represent IPTW-adjusted patient counts. P = Pembrolizumab; P + C = Pembrolizumab + Chemotherapy.

**Figure S6. Heterogeneity of chemotherapy benefit by baseline predicted survival in advanced urothelial carcinoma**

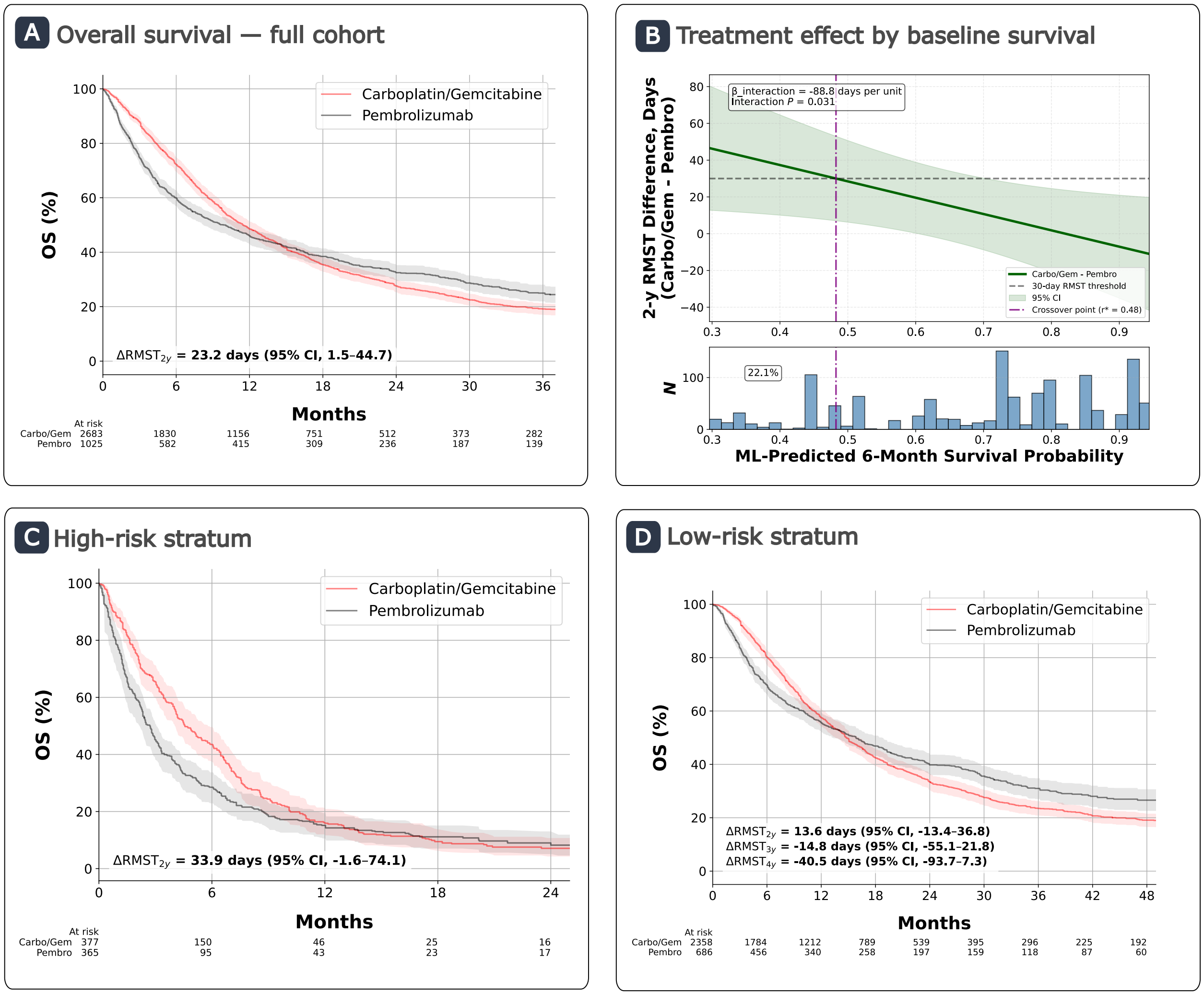

**Panel A**: IPTW-adjusted overall survival in the full cohort, demonstrating a 2-year RMST benefit with carboplatin-based chemotherapy compared with pembrolizumab monotherapy ($\Delta RMST_{2y}$ = 23.2 days, 95% CI: 1.5 to 44.7).
**Panel B**: Estimated treatment-effect function demonstrating increased benefit of carboplatin therapy as baseline predicted survival worsened ($\beta_{interaction}$ = −8.9 days per 0.1 increase in predicted 6-month survival, *P* = .03). The crossover point (r*), defined as the predicted survival probability at which combination therapy confers a ≥ 30-day improvement in 2-year RMST, occurred at r = 0.48.
**Panels C and D**: IPTW-adjusted Kaplan–Meier curves stratified by r*. Among higher-risk patients (r < 0.48), the estimated 2-year RMST difference favored carboplatin plus gemcitabine over pembrolizumab, although the 95% CI included zero ($\Delta RMST_{2y}$ = 33.9 days, 95% CI: -1.6 to 74.1). Among lower-risk patients (r ≥ 0.48), the estimated difference was smaller and also uncertain ($\Delta RMST_{2y}$ = 13.6 days, 95% CI: -13.4 to 36.8). Numbers at risk represent IPTW-adjusted patient counts rather than raw patient counts.

**Table S16. Baseline characteristics by risk stratum in recurrent/metastatic HNSCC with PD-L1 CPS ≥1 or unknown receiving pembrolizumab plus chemotherapy or pembrolizumab monotherapy**

| Characteristic | High-risk | Low-risk |
|---|---|---|
| *N* (%) | 388 (26.4) | 1,083 (73.6) |
| Predicted 6 month survival probability, median (IQR) | 0.47 (0.42-0.56) | 0.87 (0.71-0.92) |
| Age, median (IQR) | 68 (60-77) | 67 (60-74) |
| Male sex, n (%) | 286 (73.7) | 866 (80.0) |
| ECOG, n (%) | | |
| 0-1 | 168 (43.3) | 735 (67.9) |
| ≥2 | 131 (33.8) | 78 (7.2) |
| Unknown | 89 (22.9) | 270 (24.9) |
| Labs, n (%) | | |
| Creatinine >2 mg/dl | 8 (2.1) | 9 (0.8) |
| Hemoglobin <9 g/dl | 60 (15.5) | 24 (2.2) |
| WBC >11 × $10^3$/μL | 122 (31.4) | 81 (7.5) |
| ALP >200 U/L | 31 (8.0) | 14 (1.3) |
| Albumin <3 g/dl | 40 (10.3) | 11 (1.0) |
| Percent change in weight, median (IQR) | -5.0 (-10.5 to -0.7) | -0.7 (-3.4 to 1.4) |
| History of smoking, n (%) | 299 (77.1) | 821 (75.8) |
| HPV positive, n (%) | 78 (20.1) | 481 (44.4) |
| PD-L1 CPS ≥1, n (%) | 171 (44.1) | 432 (39.9) |
| Primary site, n (%) | | |
| Oropharynx | 135 (34.8) | 563 (52.0) |
| Oral cavity | 137 (35.3) | 194 (17.9) |
| Larynx/hypopharynx | 109 (28.1) | 298 (27.5) |
| Other or unknown | 7 (1.8) | 28 (2.6) |
| Sites of metastases, n (%) | | |
| Thoracic | 82 (21.1) | 233 (21.5) |
| Liver | 21 (5.4) | 35 (3.2) |
| Bone | 55 (14.2) | 88 (8.1) |
| Received pembrolizumab + chemotherapy, n (%) | 144 (37.1) | 365 (33.7) |

Patients were stratified by the survival probability crossover value (r* = 0.65), defined as the predicted 6-month survival probability at which pembrolizumab plus chemotherapy confers at least 30 days of 2-year RMST benefit over pembrolizumab monotherapy. High-risk denotes patients below this threshold; low-risk denotes patients at or above it. Percent change in weight reflects change from baseline to treatment initiation. Counts and percentages represent raw (unweighted) patient characteristics.

**Table S17. Baseline characteristics by risk stratum in advanced NSCLC with high PD-L1 (TPS ≥50%) receiving pembrolizumab plus chemotherapy or pembrolizumab monotherapy**

| Characteristic | High-risk | Low-risk |
|---|---|---|
| *N* (%) | 1,597 (28.5) | 4,008 (71.5) |
| Predicted 6 month survival probability, median (IQR) | 0.47 (0.36-0.53) | 0.83 (0.71-0.90) |
| Age, median (IQR) | 74 (66-82) | 70 (63-77) |
| Male sex, n (%) | 951 (59.5) | 1,949 (48.6) |
| ECOG, n (%) | | |
| 0-1 | 560 (35.1) | 2,529 (63.1) |
| ≥2 | 673 (42.1) | 442 (11.0) |
| Unknown | 364 (22.8) | 1,037 (25.9) |
| Labs, n (%) | | |
| Creatinine >2 mg/dl | 52 (3.3) | 25 (0.6) |
| Hemoglobin <9 g/dl | 169 (10.6) | 119 (3.0) |
| WBC >11 × $10^3$/μL | 849 (53.2) | 912 (22.8) |
| ALP >200 U/L | 184 (11.5) | 73 (1.8) |
| Albumin <3 g/dl | 311 (19.5) | 121 (3.0) |
| Percent change in weight, median (IQR) | -3.0 (-7.1 to 0.0) | -0.7 (-3.0 to 1.2) |
| History of smoking, n (%) | 1,507 (94.4) | 3,743 (93.4) |
| Histology, n (%) | | |
| Non-squamous | 1,035 (64.8) | 3,059 (76.3) |
| Squamous | 457 (28.6) | 778 (19.4) |
| NOS | 105 (6.6) | 171 (4.3) |
| Sites of metastases, n (%) | | |
| Liver | 164 (10.3) | 128 (3.2) |
| Brain | 229 (14.3) | 581 (14.5) |
| Bone | 512 (32.1) | 666 (16.6) |
| Received pembrolizumab + chemotherapy, n (%) | 435 (27.2) | 1,379 (34.4) |

Patients were stratified by the survival probability crossover value (r* = 0.59), defined as the predicted 6-month survival probability at which pembrolizumab plus chemotherapy confers at least 30 days of 2-year RMST benefit over pembrolizumab monotherapy. High-risk denotes patients below this threshold; low-risk denotes patients at or above it. Percent change in weight reflects change from baseline to treatment initiation. Counts and percentages represent raw (unweighted) patient characteristics.

**Table S18. Baseline characteristics by risk stratum in advanced urothelial carcinoma receiving carboplatin plus gemcitabine or pembrolizumab**

| Characteristic | High-risk | Low-risk |
|---|---|---|
| *N* (%) | 747 (21.5) | 2,721 (78.5) |
| Predicted 6 month survival probability, median (IQR) | 0.37 (0.30-0.48) | 0.79 (0.72-0.87) |
| Age, median (IQR) | 79 (70.5-85) | 74 (68-80) |
| Male sex, n (%) | 545 (73.0) | 1,990 (73.1) |
| ECOG, n (%) | | |
| 0-1 | 251 (33.6) | 1,567 (57.6) |
| ≥2 | 297 (39.8) | 256 (9.4) |
| Unknown | 199 (26.6) | 898 (33.0) |
| Labs, n (%) | | |
| Creatinine >2 mg/dl | 140 (18.7) | 235 (8.6) |
| Hemoglobin <9 g/dl | 163 (21.8) | 172 (6.3) |
| WBC >11 × $10^3$/μL | 363 (48.6) | 344 (12.6) |
| ALP >200 U/L | 189 (25.3) | 79 (2.9) |
| Albumin <3 g/dl | 169 (22.6) | 52 (1.9) |
| Percent change in weight, median (IQR) | -2.5 (-6.7 to 0.0) | -0.8 (-3.5 to 1.0) |
| History of smoking, n (%) | 577 (77.2) | 1,985 (73.0) |
| PD-L1 TPS ≥1%, n (%) | 30 (4.0) | 93 (3.4) |
| Sites of metastases, n (%) | | |
| Thoracic | 81 (10.8) | 256 (9.4) |
| Liver | 105 (14.1) | 112 (4.1) |
| Bone | 165 (22.1) | 278 (10.2) |
| Received pembrolizumab, n (%) | 370 (49.5) | 978 (35.9) |

Patients were stratified by the survival probability crossover value ($r^* = 0.48$), defined as the predicted 6-month survival probability at which carboplatin-based chemotherapy confers at least 30 days of 2-year RMST benefit over pembrolizumab. High-risk denotes patients below this threshold; low-risk denotes patients at or above it. Percent change in weight reflects change from baseline to treatment initiation. Counts and percentages represent raw (unweighted) patient characteristics.

**Figure S7. Heterogeneity of combination ICI benefit by baseline predicted survival in advanced BRAF-mutant melanoma**

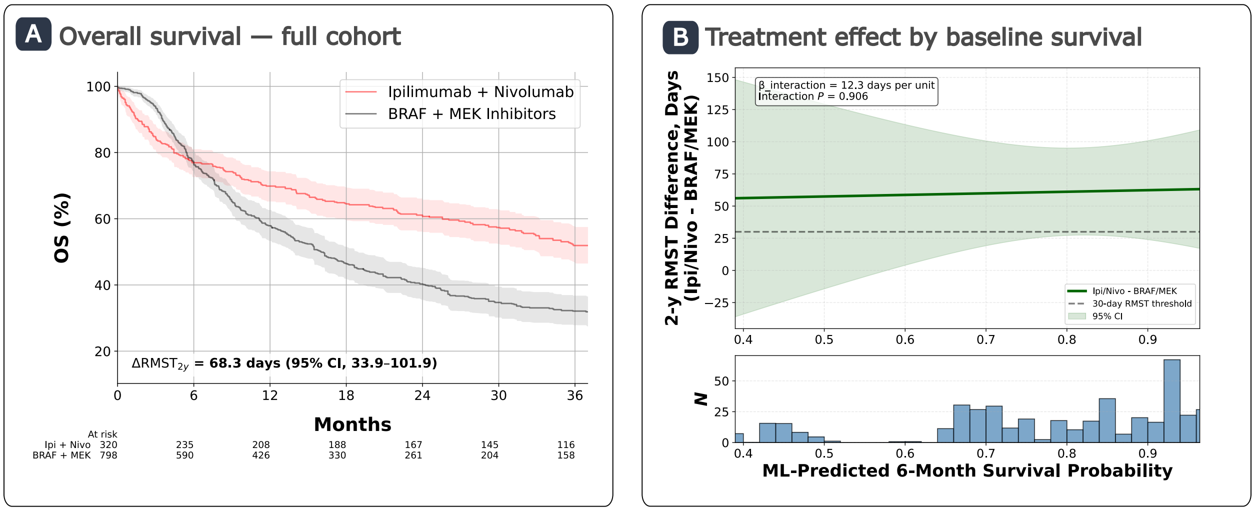

**Panel A**: IPTW-adjusted overall survival in the full cohort demonstrating improved 2-year RMST with ipilimumab/nivolumab compared with BRAF/MEK inhibitor combination therapy ($\Delta RMST_{2y}$ = 68.3 days, 95% CI: 33.9 to 101.9). Numbers at risk represent IPTW-adjusted patient counts rather than raw patient counts.

**Panel B**: Estimated treatment-effect function demonstrated no clear evidence of a linear interaction across baseline predicted survival ($\beta_{interaction}$ = 1.2 days per 0.1 increase in predicted 6-month survival; *P* = .91 ). No crossover point was identified.

#### RCT benchmarking

For study cohorts with a suitable RCT benchmark, treatment effects estimated in the full analytic cohort were compared with results from the corresponding RCT (Table S19). The HNSCC study cohort was not included in formal concordance testing because KEYNOTE-048 did not directly compare the treatment strategies evaluated here; a descriptive comparison is provided in Table S20A.

Across the four formally benchmarked comparisons, all emulations met the prespecified confidence interval and standardized-difference agreement criteria, and two met the significance agreement criterion (Table S19).

**Table S19. Concordance of target trial emulations with RCT benchmarks**

| Cohort | Trial | Treatment Comparison | RCT Estimate (95% CI) | Emulated Estimate (95% CI) | Significance Agreement | 95% CI Agreement | Stand. Diff. |
|---|---|---|---|---|---|---|---|
| NSCLC | PERSEE | Pembro + Chemo vs. Pembro | 1.3 (-0.39 to 3.0) | 0.8 (0.2 to 1.4) | No | Yes | -0.53 |
| Urothelial | KEYNOTE-361 | Pembro vs. Chemo | 0.92 (0.77-1.11) | 0.99 (0.89-1.09) | Yes | Yes | 0.68 |
| Colorectal | KEYNOTE-177 | Pembro vs. Chemo | 0.74 (0.53-1.03) | 0.59 (0.48-0.72) | No | Yes | -1.13 |
| Melanoma | DREAMseq | Ipi/Nivo vs. BRAF/MEK | 20.3% (2.6-37.9) | 20.8% (14.2-27.6) | Yes | Yes | 0.06 |

*Pembro = Pembrolizumab; Chemo = Chemotherapy; Ipi/Nivo = Ipilimumab plus Nivolumab; Stand. Diff = Standardized Difference*

Benchmarking used overall survival HRs for urothelial and colorectal cancer, the between-treatment difference in 2-year overall survival RMST (months) for NSCLC, and the absolute difference in 2-year overall survival (percentage points) for melanoma. HRs were estimated using stabilized IPTW-weighted Cox proportional hazards models. Schoenfeld residual testing suggested violation of the proportional hazards assumption in the urothelial carcinoma cohort, further supporting the use of RMST as the primary estimand in the present study.

##### Comparison with RCT by baseline predicted risk

In HNSCC, median overall survival with pembrolizumab plus chemotherapy and pembrolizumab alone in the full cohort was similar to that in the corresponding KEYNOTE-048 arms in the PD-L1 CPS ≥1 population (Table S20A). In NSCLC, the overall difference in 2-year overall survival RMST between pembrolizumab plus chemotherapy and pembrolizumab alone was similar to that reported in PERSEE. Combination therapy was associated with a larger estimated benefit among higher-risk patients, whereas the estimated benefit among lower-risk patients was smaller and not statistically significant (Table S20B). In urothelial carcinoma, lower-risk patients had median overall survival similar to that reported in KEYNOTE-361 (Table S20C). Higher-risk patients were defined as those with predicted 6-month survival below the crossover value; lower-risk patients had predicted survival at or above this value.

**Table S20 A. HNSCC: Median overall survival, months**

| Stratum | Emulated Estimate (95% CI) | KEYNOTE-048 Estimate (95% CI) |
|---|---:|---:|
| Overall | 14.1 (12.9-16.6) vs. 13.6 (11.7-17.1) | 13.6 (10.7-15.5) vs. 12.3 (10.8-14.8) |
| High-risk | 7.5 (6.0-9.4) vs. 3.7 (3.1-4.8) | — |
| Low-risk | 18.4 (14.6-21.1) vs. 20.4 (17.9-22.5) | — |

*Pembrolizumab plus chemotherapy is listed first. KEYNOTE-048 did not formally compare these arms; estimates from CPS ≥1 cohort are shown for context only.*

**Table S20 B. NSCLC: Between-treatment difference in 2-year overall survival RMST, months**

| Stratum | Emulated Estimate (95% CI) | PERSEE Estimate (95% CI) |
|---|---:|---:|
| Overall | 0.8 (0.2 to 1.4) | 1.3 (-0.39 to 3.0) |
| High-risk | 1.2 (0.1 to 2.3) | — |
| Low-risk | 0.5 (-0.1 to 1.1) | — |

*Positive differences favor pembrolizumab plus chemotherapy over pembrolizumab alone.*

**Table S20 C. Urothelial: Median overall survival, months**

| Stratum | Emulated Estimate (95% CI) | KEYNOTE-361 Estimate (95% CI) |
|---|---:|---:|
| Overall | 10.1 (8.2 to 11.4) vs. 11.5 (10.5 to 12.6) | 15.6 (12.1-17.9) vs.14.3 (12.3-16.7) |
| High-risk | 2.7 (2.2-3.1) vs. 4.6 (4.0-5.8) | — |
| Low-risk | 15.9 (13.3-18.8) vs. 14.8 (13.6-16.2) | — |

*Pembrolizumab is listed first and chemotherapy second: carboplatin plus gemcitabine in the real-world cohort and platinum-based chemotherapy in KEYNOTE-361.*

Higher-risk patients were more frequently classified as ineligible for the corresponding RCT based on ECOG performance status or laboratory criteria: 22.4% versus 4.9% in HNSCC, 26.2% versus 5.1% in NSCLC, and 56.0% versus 21.1% in urothelial carcinoma (Table S21). These findings suggest that risk stratification identified clinically vulnerable patients less well represented in the trials, although eligibility was indeterminate for a substantial proportion of each cohort.

**Table S21. Estimated RCT eligibility by baseline predicted risk in the real-world HNSCC, NSCLC, and urothelial cohorts**

| Cohort | Real-World Stratum | Eligible, *N* (%) | Ineligible, *N* (%) | Indeterminate, *N* (%) |
|---|---|---|---|---|
| HNSCC | Overall | 758 (51.5) | 140 (9.5) | 573 (39.0) |
| | High-risk | 178 (45.9) | 87 (22.4) | 123 (31.7) |
| | Low-risk | 580 (53.6) | 53 (4.9) | 450 (41.6) |
| NSCLC | Overall | 2,826 (50.4) | 623 (11.1) | 2,156 (38.5) |
| | High-risk | 704 (44.1) | 419 (26.2) | 474 (29.7) |
| | Low-risk | 2,122 (52.9) | 204 (5.1) | 1,682 (42.0) |
| Urothelial | Overall | 1,223 (35.3) | 992 (28.6) | 1,253 (36.1) |
| | High-risk | 175 (23.4) | 418 (56.0) | 154 (20.6) |
| | Low-risk | 1,048 (38.5) | 574 (21.1) | 1,099 (40.4) |

Patients were classified as eligible when ECOG performance status and all required laboratory criteria were met, ineligible when at least one criterion was violated, and indeterminate when no violation was identified but at least one required variable was missing. Categories are mutually exclusive and sum to the full cohort.

##### RCC concordance analysis

In the RCC concordance cohort, the estimated benefit of ipilimumab plus nivolumab relative to single-agent VEGF-targeted therapy increased as baseline predicted survival decreased (Table S22). The estimated 3-year RMST benefit was 84.2 days at the 10th percentile and 1.2 days at the 90th percentile of predicted survival, a difference of 83.0 days (95% CI, 14.8 to 151.2). The treatment-effect curve crossed the 30-day RMST benefit threshold at a calibrated 6-month predicted survival probability of 0.81. Twelve-month overall survival was substantially lower in the higher-risk than lower-risk stratum with both ipilimumab plus nivolumab (48.5% vs. 82.1%) and single-agent VEGF-targeted therapy (42.9% vs. 80.5%; Table S22, Panel B).

**Table S22 A. Continuous treatment-effect analysis**

| Comparison | Overall ΔRMST, days | TE at 10th percentile, days | TE at 90th percentile, days | 10th–90th difference, days | Interaction *P* |
|---|---|---|---|---|---|
| Ipi/Nivo vs. single-agent VEGF-TKI | 28.8 (-4.8 to 62.1) | 84.2 (29.9 to 138.4) | 1.2 (-38.2 to 40.6) | 83.0 (14.8 to 151.2) | .02 |

**Table S22 B. Risk-stratified RMST differences and 12-month overall survival**

| Baseline predicted 6-month survival | Stratum | ΔRMST, days | 12-month OS: Ipi/Nivo | 12-month OS: VEGF-TKI |
|---|---|---|---|---|
| r < 0.81 | High-risk | 44.5 (-7.9 to 97.1) | 48.5% (42.9 to 54.7) | 42.9% (39.8 to 45.8) |
| r ≥ 0.81 | Low-risk | 19.3 (-21.9 to 58.1) | 82.1% (78.2 to 85.5) | 80.5% (78.4 to 82.3) |

Treatment-effect heterogeneity in the RCC concordance cohort is shown in Figure S8, and baseline characteristics by risk stratum are shown in Table S23.

**Figure S8. Heterogeneity of combination ICI benefit by baseline predicted survival in advanced RCC**

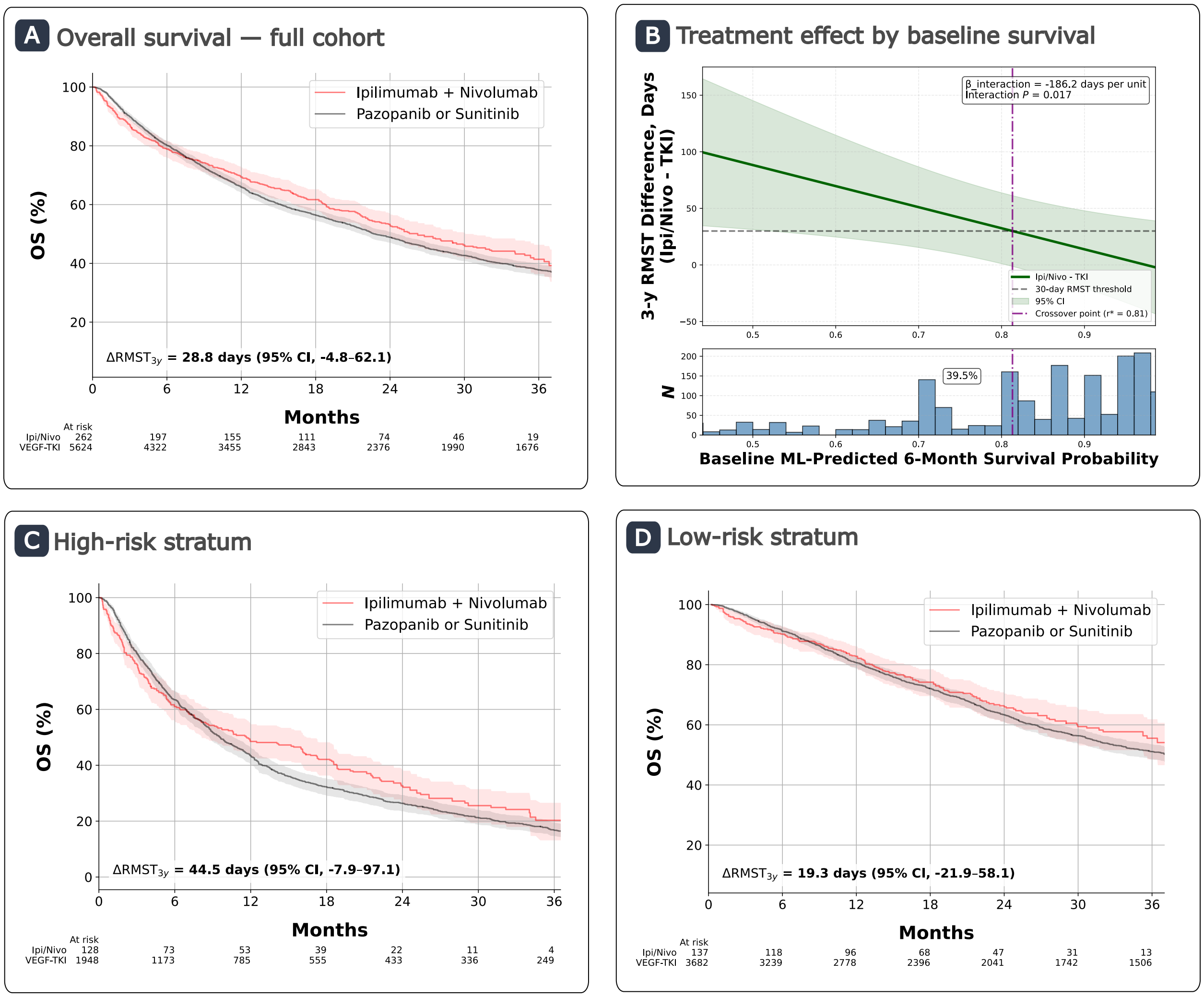

**Panel A**: IPTW-adjusted overall survival in the full cohort, with no meaningful difference in 3-year RMST between ipilimumab/nivolumab and single-agent anti-angiogenic therapy ($\Delta RMST_{3y}$ = 28.8 days, 95% CI: -4.8 to 62.1).

**Panel B**: Estimated treatment-effect function demonstrating increased benefit of ipilimumab/nivolumab therapy as baseline predicted survival worsened ($\beta_{interaction}$ = −18.6 days per 0.1 increase in predicted 6-month survival, *P* = .02). The crossover point (r*), defined as the predicted survival probability at which ipilimumab/nivolumab therapy confers a ≥ 30-day improvement in 3-year RMST, occurred at r = 0.81.

**Panels C and D**: IPTW-adjusted Kaplan–Meier curves stratified by r*. Among higher-risk patients (r < 0.81), ipilimumab/nivolumab therapy was associated with a higher RMST ($\Delta RMST_{3y}$ = 44.5 days, 95% CI: -7.9 to 97.1). Among lower-risk patients (r ≥ 0.81), no meaningful difference was observed ($\Delta RMST_{3y}$ = 19.3 days, 95% CI: -21.9 to 58.1). Numbers at risk represent IPTW-adjusted patient counts rather than raw patient counts.

**Table S23. Baseline characteristics by risk stratum in metastatic clear cell RCC receiving ipilimumab/nivolumab or single-agent antiangiogenic therapy**

| Characteristic | High-risk | Low-risk |
|---|---|---|
| *N* (%) | 1,540 (37.1) | 2,610 (62.9) |
| Predicted 6 month survival probability, median (IQR) | 0.70 (0.53-0.73) | 0.93(0.87-0.96) |
| Age, median (IQR) | 69 (61-76) | 65.5 (58-72) |
| Male sex, *N* (%) | 1,041 (67.6) | 1,881 (72.1) |
| ECOG, *N* (%) | | |
| 0-1 | 574 (37.3) | 1,201 (46.0) |
| ≥2 | 318 (20.6) | 98 (3.8) |
| Unknown | 648 (42.1) | 1,311 (50.2) |
| IMDC risk, *N* (%) | | |
| Favorable | 61 (4.0) | 1,046 (40.1) |
| Intermediate/poor | 1,479 (96.0) | 1,564 (59.9) |
| Labs, *N* (%) | | |
| Creatinine >2 mg/dl | 126 (8.2) | 93 (3.6) |
| Hemoglobin <12 g/dl | 846 (54.9) | 481 (18.4) |
| Calcium > 10.2 mg/dl | 142 (9.2) | 117 (4.5) |
| Neutrophil >7 × $10^3$/μL | 358 (23.2) | 109 (4.2) |
| Platelets > 400,000 cells/μL | 325 (21.1) | 148 (5.7) |
| ALP >200 U/L | 154 (10.0) | 23 (0.9) |
| Albumin <3 g/dl | 181 (11.8) | 12 (0.5) |
| Percent change in weight, median (IQR) | -2.0 (-5.9 to 0.0) | -0.1 (-2.2 to 1.5) |
| Sites of metastases, *N* (%) | | |
| Thoracic | 283 (18.4) | 554 (21.2) |
| Liver | 76 (4.9) | 91 (3.5) |
| Bone | 373 (24.2) | 419 (16.1) |
| CNS | 95 (6.2) | 88 (3.4) |
| <1 year from time of diagnosis to systemic therapy, *N* (%) | 1,266 (82.2) | 1,177 (45.1) |
| Received ipilimumab/nivolumab, *N* (%) | 316 (20.5) | 425 (16.3) |

Patients were stratified by the survival probability crossover value (r* = 0.81), defined as the predicted 6-month survival probability at which ipilimumab/nivolumab confers at least 30 days of 3-year RMST benefit over single-agent antiangiogenic therapy. High-risk denotes patients below this threshold; low-risk denotes patients at or above it. Percent change in weight reflects change from baseline to treatment initiation. Counts and percentages represent raw (unweighted) patient characteristics.

#### Sensitivity analyses

*Biomarker result availability at treatment initiation*

Restricting biomarker eligibility to results available on or before the index date reduced cohort size in the three studies defined by a positive biomarker result: NSCLC decreased by 8.9%, colorectal cancer by 10.5%, and melanoma by 16.8% (Table S24). In contrast, urothelial carcinoma had no biomarker-based eligibility requirement, and its cohort size was unchanged. The HNSCC cohort, which permitted PD-L1 CPS ≥1 or unknown, increased modestly by 1.8% because patients with unknown status at treatment initiation were eligible even if subsequent results showed CPS <1. Therefore, in HNSCC and urothelial carcinoma, the timing restriction primarily affected the biomarker information available to the prognostic model rather than cohort eligibility.

The overall treatment-effect patterns were preserved under the restricted timing window. In HNSCC and urothelial carcinoma, the treatment-by-risk interactions remained statistically significant, with similar 10th–90th percentile treatment-effect differences and crossover values. In NSCLC, the estimated 10th–90th percentile treatment-effect difference increased from 35.3 to 46.3 days and the interaction reached statistical significance (P = .04), with a modest shift in the crossover value (0.59 to 0.66). In colorectal cancer and melanoma, there remained no statistically significant treatment-by-risk interaction or crossing of the prespecified 30-day RMST threshold.

**Table S24. Sensitivity of treatment-effect estimates to biomarker result availability**

| | Results available by day +30 | | | | Result available by index date | | | |
|---|---|---|---|---|---|---|---|---|
| **Cohort** | ***N*** | **10-90th TE difference, days** | ***P*** | **r*** | ***N*** | **10-90th TE difference, days** | ***P*** | **r*** |
| HNSCC | 1,471 | 96.7 (33.7 to 159.7) | .003 | 0.65 | 1,498 | 108.0 (38.0 to 178.0) | .002 | 0.67 |
| NSCLC | 5,605 | 35.3 (-3.2 to 73.8) | .07 | 0.59 | 5,104 | 46.3 (2.6 to 89.9) | .04 | 0.66 |
| Urothelial | 3,468 | 51.4 (4.8 to 98.1) | .03 | 0.48 | 3,468 | 53.3 (8.9 to 98.2) | .02 | 0.50 |
| Colorectal | 1,043 | 27.2 (-25.9 to 80.2) | .32 | — | 933 | 44.5 (-10.4 to 99.3) | .11 | — |
| Melanoma | 1,069 | -6.2 (-108.8 to 96.5) | .91 | — | 889 | 13.8 (-117.4 to 145.0) | .84 | — |

Across Tables S24–S28, S30, and S32, the 10th–90th treatment-effect (TE) difference is the estimated RMST difference at the overlap-weighted 10th percentile of calibrated 6-month survival probability minus that at the 90th percentile, in days; parentheses give 95% CIs. *P* values test treatment-by-risk interaction. $r^*$ is the calibrated 6-month survival probability at which the estimated TE reaches the prespecified 30-day RMST benefit threshold; dash indicates that no threshold crossing was identified within the evaluated risk range. Six-month AUC was estimated in held-out test data where reported.

*Alternative prognostic model*

Replacing the gradient-boosted prognostic model with a ridge-penalized Cox model modestly reduced held-out 6-month discrimination across cohorts (Table S25). The estimated treatment-effect differences between the 10th and 90th percentiles of predicted survival were directionally consistent across models, and the strength of evidence for treatment-by-risk interaction was largely unchanged. Estimated $r^*$ values were similar in HNSCC (0.65 vs 0.67) and NSCLC (0.59 vs 0.62) but shifted from 0.48 to 0.61 in urothelial carcinoma. Neither model identified a threshold in colorectal cancer or melanoma.

**Table S25. Comparing treatment-by-risk interaction estimates from gradient-boosted and ridge-penalized Cox models**

| | Gradient-Boosted Model | | | | Penalized Cox Model | | | |
|---|---|---|---|---|---|---|---|---|
| **Cohort** | **6-mo AUC** | **10-90th TE difference, days** | ***P*** | **r*** | **6-mo AUC** | **10-90th TE difference, days** | ***P*** | **r*** |
| HNSCC | 0.751 | 96.7 (33.7 to 159.7) | .003 | 0.65 | 0.741 | 101.9 (31.1 to 172.8) | .005 | 0.67 |
| NSCLC | 0.773 | 35.3 (-3.2 to 73.8) | .07 | 0.59 | 0.737 | 45.3 (-0.1 to 90.6) | .05 | 0.62 |
| Urothelial | 0.796 | 51.4 (4.8 to 98.1) | .03 | 0.48 | 0.776 | 55.3 (5.1 to 105.4) | .03 | 0.61 |
| Colorectal | 0.791 | 27.2 (-25.9 to 80.2) | .32 | — | 0.759 | 49.3 (-40.9 to 139.6) | .28 | — |
| Melanoma | 0.798 | -6.2 (-108.8 to 96.5) | .91 | — | 0.786 | -7.3 (-99.2 to 84.7) | .88 | — |

*Alternative prognostic model-development population*

To assess whether associations between baseline features and treatment selection in the full model-development population influenced the predicted baseline survival, models were retrained using only chemotherapy-treated patients. Retraining the prognostic model using only chemotherapy-treated patients yielded similar 6-month discrimination in HNSCC and colorectal cancer test sets (Table S26). In HNSCC, the 10th–90th TE difference remained substantial (96.7 versus 88.8 days), with similar $r^*$values (0.65 versus 0.66). In colorectal cancer, neither prognostic model identified a threshold crossing, and the treatment-by-risk interaction was nonsignificant under both specifications. These findings did not materially change the interpretation of either cohort.

**Table S26. Treatment-effect heterogeneity under all-patient and single-arm prognostic model specifications**

| | All-patient prognostic model | | | | Single-arm prognostic model | | | |
|---|---|---|---|---|---|---|---|---|
| **Cohort** | **6-mo AUC** | **10-90th TE difference, days** | ***P*** | **r*** | **6-mo AUC** | **10-90th TE difference, days** | ***P*** | **r*** |
| HNSCC | 0.751 | 96.7 (33.7 to 159.7) | .003 | 0.65 | 0.755 | 88.8 (24.4 to 153.2) | .007 | 0.66 |
| Colorectal | 0.791 | 27.2 (-25.9 to 80.2) | .32 | — | 0.798 | 59.1 (-23.4 to 141.7) | .16 | — |

*Alternative functional form*

Allowing the treatment-effect function to vary through a natural cubic spline yielded similar qualitative findings for HNSCC, NSCLC, urothelial carcinoma, and melanoma (Table S27; Figure S9). In HNSCC, NSCLC, and urothelial carcinoma, the estimated 30-day threshold crossings differed by no more than 0.10 in predicted 6-month survival probability between specifications. Neither model identified a threshold crossing in colorectal cancer or melanoma.

Colorectal cancer showed a significant global spline interaction (*P* = .001), with an inverted U-shaped treatment-effect curve suggesting that the estimated benefit of pembrolizumab was greatest at intermediate predicted survival probabilities. Nevertheless, the estimated benefit remained above the prespecified 30-day RMST threshold across the plotted risk range. Thus, the spline suggested variation in the magnitude of benefit, but did not identify a risk range in which chemotherapy became the favored strategy.

We retained the linear model as the primary analysis because it provides a parsimonious, readily interpretable summary of the treatment-effect relationship and has shown favorable performance relative to more flexible specifications in simulation studies, particularly at moderate sample sizes [9].

**Table S27. Comparison of linear and spline treatment-effect estimates**

| | Linear model | | | Spline model | | |
|---|---|---|---|---|---|---|
| **Cohort** | **$10\text{-}90^{th}$ TE difference, days** | **Interaction *P*** | **r*** | **$10\text{-}90^{th}$ TE difference, days** | **Global *P*** | **r*** |
| HNSCC | 96.7 (33.7 to 159.7) | .003 | 0.65 | 123.6 (55.9 to 191.4) | .001 | 0.75 |
| NSCLC | 35.3 (-3.2 to 73.8) | .07 | 0.59 | 30.2 (-9.2 to 69.5) | .09 | 0.55 |
| Urothelial | 51.4 (4.8 to 98.1) | .03 | 0.48 | 45.9 (-2.3 to 94.1 ) | .05 | 0.45 |
| Colorectal | 27.2 (-25.9 to 80.2) | .32 | — | 114.4 (32.4 to 196.4) | .001 | — |
| Melanoma | -6.2 (-108.8 to 96.5) | .91 | — | -18.5 (-116.3 to 79.3) | .73 | — |

#### Figure S9. Comparing linear and natural cubic spline treatment effect function across study cohorts

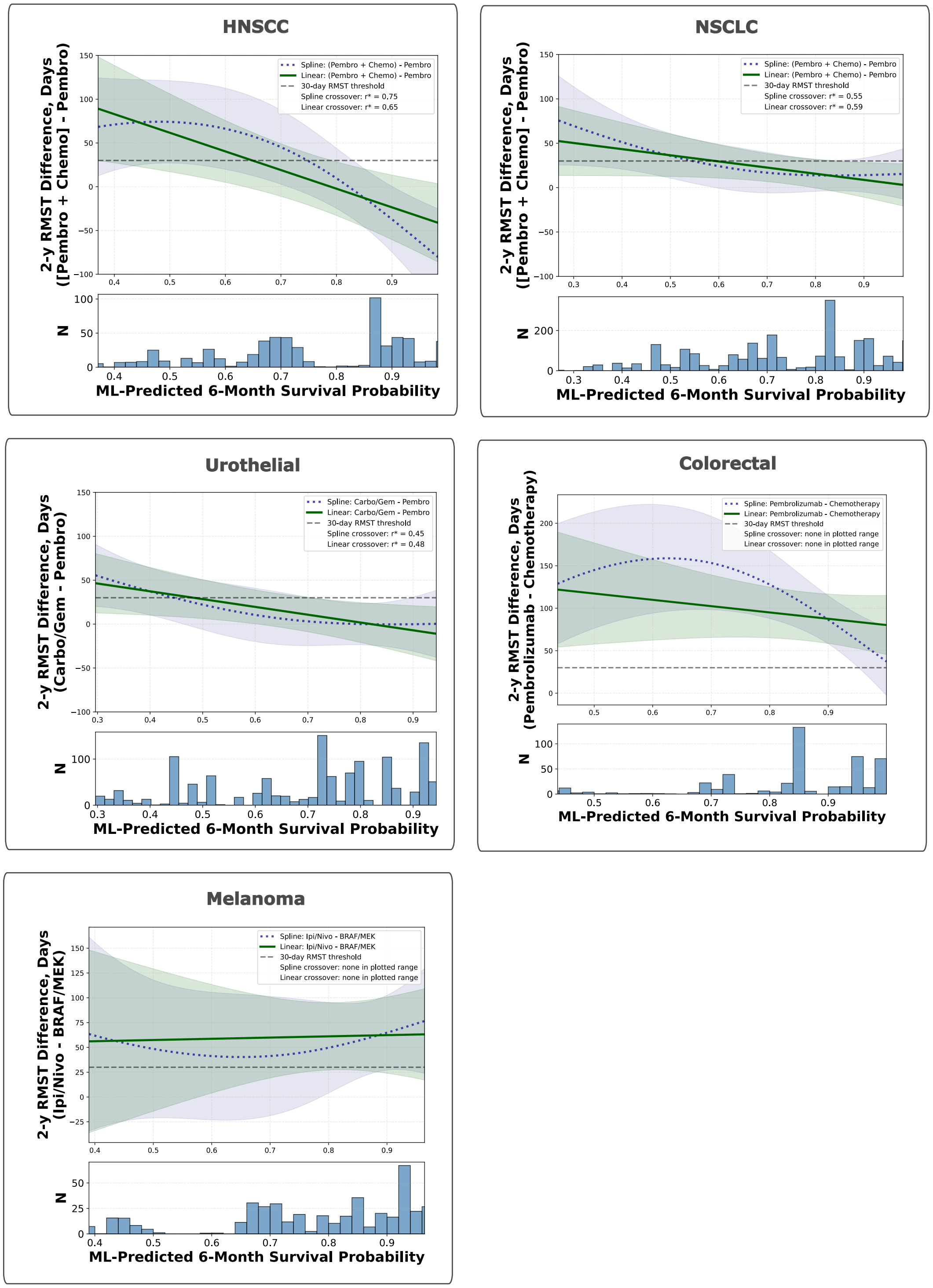

Each panel shows the estimated treatment-effect function TE(r), the difference in 2-year RMST between treatment arms as a function of calibrated 6-month survival probability, from the primary linear specification (solid green line) and a natural cubic spline (dotted blue line, df=3), with 95% confidence intervals. The dashed horizontal line indicates 30-day RMST threshold. For cohorts where a 30-day crossover point was identified, r* values for each specification are reported in the legend. No crossover threshold was identified for dMMR/MSI-H colorectal cancer or BRAF-mutant melanoma under the primary linear or spline specification. Histograms show the overlap-weighted distribution of calibrated 6-month survival probability.

*Alternative RMST horizon*

At a 1-year rather than 2-year RMST horizon, the 10th–90th TE differences remained positive in HNSCC, NSCLC, and urothelial carcinoma, and the linear interaction tests were significant in all three cohorts (Table S28). The absolute differences were smaller at 1 year, as expected with a shorter accumulation period. Colorectal cancer and melanoma had nonsignificant linear interactions at both horizons.

The 1-year colorectal analysis yielded an estimated $r^*$ of 0.61, whereas the 2-year analysis had no threshold crossing (Table S28). At 1 year, however, the estimated pembrolizumab–chemotherapy benefit varied by only 11.2 days between the 10th and 90th percentiles of predicted survival (95% CI, −15.0 to 37.3), with no significant treatment-by-risk interaction. The threshold crossing therefore did not suggest a clinically meaningful change in treatment preference across baseline risk.

**Table S28. Treatment-by-risk interaction at 1- and 2-year RMST horizons**

| | 1-yr RMST | | | 2-yr RMST | | |
|---|---|---|---|---|---|---|
| **Cohort** | **10-90$^{th}$ TE difference, days** | ***P*** | **r*** | **10-90$^{th}$ TE difference, days** | ***P*** | **r*** |
| HNSCC | 48.3 (18.2 to 78.3) | .002 | 0.57 | 96.7 (33.7 to 159.7) | .003 | 0.65 |
| NSCLC | 27.6 (8.9 to 46.3) | .004 | 0.42 | 35.3 (-3.2 to 73.8) | .07 | 0.59 |
| Urothelial | 28.6 (6.4 to 50.7) | .01 | 0.59 | 51.4 (4.8 to 98.1) | .03 | 0.48 |
| Colorectal | 11.2 (-15.0 to 37.3) | .40 | 0.61 | 27.2 (-25.9 to 80.2) | .32 | — |
| Melanoma | -12.7 (-60.9 to 35.5) | .61 | — | -6.2 (-108.8 to 96.5) | .91 | — |

*Alternative clinically meaningful benefit thresholds*

Varying the clinically meaningful RMST benefit threshold changed the predicted 6-month survival probability at which the treatment-effect function reached that benefit threshold (the crossover value), and therefore changed the size of the resulting risk strata (Table S29). Lower RMST benefit thresholds generally produced higher survival probability crossover values and classified a larger proportion of patients as higher risk, whereas higher RMST benefit thresholds produced lower crossover values and smaller higher-risk strata.

In HNSCC, the treatment-effect function reached all evaluated RMST benefit thresholds within the observed 6 month survival probabilities. In NSCLC and urothelial carcinoma, it also reached all evaluated RMST benefit thresholds, but classified fewer than 1% of patients as higher risk at the 60-day threshold. In colorectal cancer, estimated pembrolizumab benefit remained above all evaluated thresholds across the observed range. In melanoma, the treatment-effect function crossed the 60-day RMST benefit threshold only at the lower end of the predicted survival distribution, while remaining above 60 days across most of the spectrum, consistent with a large overall treatment benefit rather than meaningful treatment-effect heterogeneity.

**Table S29. Crossover value by RMST threshold**

| Cohort | 14-day r* (% High-risk) | 30-day r* (% High-risk) | 60-day r* (% High-risk) |
|---|---|---|---|
| HNSCC | 0.72 (48.9) | 0.65 (26.4) | 0.51 (15.1) |
| NSCLC | 0.82 (58.7) | 0.59 (28.5) | 0.16 (<1) |
| Urothelial | 0.66 (37.9) | 0.48 (21.5) | 0.14 (<1) |
| Colorectal | — | — | — |
| Melanoma | — | — | 0.71 (33.4) |

*Analysis restricted to PD-L1 CPS ≥1 in HNSCC*

Among the 603 patients with documented PD-L1 CPS ≥1, the treatment-by-risk pattern was similar in direction and magnitude to that in the full HNSCC cohort, with a comparable 30-day benefit threshold, although estimates were less precise (Table S30).

**Table S30. Treatment-effect heterogeneity in the HNSCC cohort restricted to PD-L1 CPS ≥1**

| PD-L1 CPS ≥1 or Unknown | | | | PD-L1 CPS ≥1 | | | |
|---|---|---|---|---|---|---|---|
| *N* | 10-90th TE difference, days | *P* | r* | *N* | 10-90th TE difference, days | *P* | r* |
| 1471 | 96.7 (33.7 to 159.7) | .003 | 0.65 | 603 | 112.7 (7.1 to 218.4) | .04 | 0.61 |

#### Exploratory Analyses

*Subsequent therapies and censoring at ICI initiation*

Subsequent ICI use was common in the urothelial and colorectal cancer chemotherapy arms (Table S31), motivating the exploratory censor-at-switch analyses. Percentages for the most common subsequent therapy are calculated among patients who received second-line treatment.

**Table S31. Subsequent therapies following first-line treatment**

| Cohort | First-line treatment | Received subsequent systemic therapy, % | Most common subsequent therapy (%) |
|---|---|---|---|
| HNSCC | Pembrolizumab + Chemotherapy | 38.7 | Cetuximab (13.7) |
| | Pembrolizumab | 40.6 | Cetuximab (14.3) |
| NSCLC | Pembrolizumab + Chemotherapy | 35.0 | Docetaxel + ramucirumab (15.4) |
| | Pembrolizumab | 30.1 | Carboplatin + pemetrexed + pembrolizumab (14.4) |
| Urothelial | Carboplatin + Gemcitabine† | 54.3 | Pembrolizumab (25.7) |
| | Pembrolizumab | 26.7 | Enfortumab vedotin (32.2) |
| Colorectal | Pembrolizumab | 27.9 | Ipilimumab + nivolumab (12.6) |
| | Chemotherapy | 62.8 | Pembrolizumab (33.3) |
| Melanoma | Ipilimumab + nivolumab | 40.0 | Dabrafenib + trametinib (41.9) |
| | BRAF/MEK | 52.0 | Ipilimumab + nivolumab (29.8) |

†Maintenance avelumab following first-line carboplatin was considered continuation maintenance rather than subsequent-line therapy and is therefore not included in this table.

Censoring chemotherapy-treated patients at initiation of second-line ICI therapy preserved the qualitative treatment-by-risk patterns (Table S32). In urothelial carcinoma, the 10th–90th percentile treatment-effect contrast increased from 51.4 to 104.1 days, and the interaction remained evident ($P < .001$). The estimated 30-day RMST benefit threshold shifted from 0.48 to 0.67. In colorectal cancer, there remained no clear evidence of a treatment-by-risk interaction ($P = .89$).

**Table S32. Treatment-effect heterogeneity after censoring at subsequent ICI initiation**

| | Primary analysis | | | Censoring at next ICI initiation | | |
|---|---|---|---|---|---|---|
| Cohort | 10-90th TE difference, days | p | r* | 10-90th TE difference, days | p | r* |
| Urothelial | 51.4 (4.8 to 98.1) | .03 | 0.48 | 104.1 (52.7 to 155.4) | <.001 | 0.67 |
| Colorectal | 27.2 (-25.9 to 80.2) | .32 | — | 4.0 (-51.4 to 59.5) | .89 | — |

These findings should be interpreted cautiously. Patients receiving subsequent ICI must survive long enough and remain fit enough to receive additional therapy, whereas patients who never switch include both those who die early and those with durable benefit from first-line treatment. Censoring at switching may therefore introduce informative censoring, with an uncertain direction of bias.
